# Research In Your Mailbox: Remote Blood Sampling Enables Longitudinal Studies in Underserved Groups

**DOI:** 10.64898/2026.02.05.26345688

**Authors:** Filip Stefanovic, Ingrid Robertson, Kathleen Moloney, Jane Edelson, Serena Nguyen, Victoria Shinkawa, Keila Uchimura, Angela Lin, Lan Le, Jodie C. Tokihiro, Meg G. Takezawa, David Phan, Joshua T. Schiffer, Michael Boeckh, Karen N. Adams, Alpana Waghmare, Nicole A. Errett, Erwin Berthier, Fang Yun Lim, Ashleigh B. Theberge

**Affiliations:** Department of Chemistry, University of Washington, Seattle, WA, USA; Department of Environmental and Occupational Health Sciences, University of Washington, Seattle, WA, USA; Department of Medicine, University of Washington, Seattle, WA, USA; Department of Chemistry, University of Hawaii at Hilo, Hilo, HI 96720; Vaccine and Infectious Diseases Division, Fred Hutchinson Cancer Center, Seattle, WA, USA; Clinical Research Division, Fred Hutchinson Cancer Center, Seattle, WA, USA; Department of Pediatrics, Division of Infectious Diseases, University of Washington, Seattle, WA, USA; Center for Clinical and Translational Research, Seattle Children’s Research Institute, Seattle, WA, USA; Department of Urology, University of Washington School of Medicine, Seattle, WA, USA

## Abstract

**Background:** When leveraged for longitudinal studies, remote sampling with transcriptomic readouts is a powerful tool for studying host immune response. Remote study flexibility circumvents common barriers to research participation including transportation and scheduling. We investigated the effectiveness of a remote design for reaching women from underrepresented, underserved, and underreported (U3) populations per the NIH definition.

**Methods:** U3 women positive for COVID-19 were tracked over the course of 6 months. Participants self-collected nasal and homeRNA-stabilized blood samples. homeRNA is a platform for remote self-collection of blood samples with RNA stabilization. Five samplings were completed in 25 days (month 1). A subset of participants likely to develop post-acute sequelae of COVID-19 (PASC) and their age-matched controls were selected for extended sampling (month 3). All participants were resurveyed at months 4, 5, and 6 about their symptoms.

**Results:** Forty participants were fully enrolled in the study, and 39 completed all study components. Survey responses show high satisfaction in the study, where all participants who completed the study (*n* = 39/39) indicated they would be willing to participate in a similar study again, with most (*n* = 32/39) indicating a willingness to participate for up to 4 years with around 15 samples collected per year.

**Conclusions:** The high participant retention (*n* = 39/40) and satisfaction in this study indicates the utility of a remote study design for longitudinal research. We also find that study topic, flexibility of sampling, and positive interactions with the study team are important factors for participant recruitment and retention.

**Summary:** 40 U3 women who tested positive for SARS-CoV-2 were tracked over 6 months with blood self-sampling. High retention and positive participant experience suggest that the increased flexibility of a fully remote design facilitates participation of individuals who may otherwise be excluded from clinical research.

## Introduction

Remote research has the potential to remove obstacles to participation that have resulted in an underrepresentation of certain groups of people. Specifically, the remote study model allows for greater flexibility with sampling time and location, which facilitates participation of individuals with challenging schedules, those who are caregivers, as well as those who have limited access to transportation.^1,2^ While some barriers to participation such as awareness of ongoing research and mistrust in science and medicine^3^ cannot be easily overcome, online recruitment and the ability to participate from home reduce the discomfort of going to a medical facility.^4^ Bolstering the participation of underreported groups, researchers can build more robust datasets and develop medical treatment and interventions that are effective for everyone.^5^

Moreover, remote studies have the potential to afford opportunities for sampling at relevant times and frequencies, especially with sensitive molecular signatures associated with transient events, such as wildfire smoke exposure, infectious disease, and response to medical treatment and interventions. By eliminating the need for in-person visits, remote research also limits risk of infection for both the participant and the research staff. In the wake of the COVID-19 pandemic, self-sampling allowed continuation of work despite closures of large research centers.^6,7^ Remote research also allows for a single study team to reach populations across the entire country under one Institutional Review Board (IRB) protocol.

Longitudinal sampling with high temporal frequency is a powerful tool for elucidating underlying mechanisms of disease progression.^8,9^ To this end, our lab previously developed an at-home blood sampling technology called homeRNA. The homeRNA platform allows participants to self-collect their blood and stabilize the RNA.^10^ We have already demonstrated the utility of homeRNA in remote longitudinal studies in respiratory infections and have been able to sequence and interpret data from the resulting stabilized blood samples.^10–15^

In the work herein, we present a homeRNA-based longitudinal study tracking the progression of the host response to the SARS-CoV-2 virus in women defined as underrepresented, underserved, and underreported (U3) according to the NIH definition. We chose post-acute sequelae of COVID-19 (PASC), more commonly known as long COVID, as a case study for implementing homeRNA in U3 populations participating in longitudinal studies.

PASC is described as a condition with symptoms resulting from the COVID-19 infection and lasting for weeks, months, or even years following original infection.^16^ Since the onset of the COVID-19 pandemic, studies have been published proposing mechanism and health implications of PASC.^17–20^ While significant work has been done to better understand this condition, at the time of writing, a standard definition is still evolving.^21–23^

To contribute to the emerging classification of infection blood biomarkers of PASC, we recruited a cohort of 40 COVID-19 positive women and tracked their symptoms over the course of 6 months. During the first month of infection, the participants collected blood samples with homeRNA, as well as nasal swabs. Given that we previously demonstrated the ability to detect and monitor pre-symptomatic signatures of COVID-19, one of the main objectives of the present study design was to capture early infection blood biomarkers to better understand potential pathways that are involved in the progression to PASC.^12,14^

Here, we assess elements of longitudinal studies and the impacts of specific study design choices on accessibility and overall study experience. Our data provide insight into effective strategies for high participant satisfaction, offering a roadmap for future study design enabling effective recruitment and retention of U3 participants.

## Methods

### Inclusion and Exclusion Criteria

Participants were recruited under a protocol approved by the University of Washington Institutional Review Board study number STUDY00016154. To meet the inclusion criteria subject had to be (1) aged 18 and older, (2) self-reported positive COVID-19 test result within 7 days of screen date, and (3) a woman of understudied, underrepresented, and underreported populations (U3). U3 is defined by NIH to include African American or Black, Hispanic or Latino, American Indian or Alaska Native, Asian American, Native Hawaiian and Other Pacific Islander populations, socioeconomically disadvantaged populations (see **Supplement 1** for NIH Disadvantaged Background criteria), underserved rural populations, and sexual and gender minorities. Individuals who (1) were unable to give consent, (2) were pregnant individuals by self-report, (3) were categorized as other protected populations (children, prisoners), or (4) did not complete the initial enrollment survey within 48 hours of consent were excluded from the study. These criteria were guided by the stated research goals of our funding source, the U3 Supplement mechanism of NIH Office of Research on Women’s Health.

### Survey Design

All surveys were built and collected using the University of Washington (UW) secure ITHS REDCap platform. These included: (1) screening surveys, (2) informed consent (ICF), (3) health history, (4) shipping information, (5) sampling surveys, (6) closing surveys, and (7) monthly follow-up surveys. The survey questions were formulated using questions from previous studies^12–14^, the PhenX directory, and the Consolidated Framework for Implementation Research (CFIR). The complete surveys are included in the **Supplement 2**.

### Recruiting and Study Initiation

Participants were recruited through advertising between March and June of 2023. This was primarily done through Facebook and the help of an advertising service, WATE TV (for a sample ad and study timeline, see **Figures e1 and e2**). Participants who responded to the ads were screened for the inclusion and exclusion criteria and were balanced for age, race/ethnicity, and state of residence in an effort to capture a diverse cohort that closely corresponds to the demographics of the United States. After the screening process, prospective participants were sent the informed consent form (ICF) found in **Supplement 2**. Upon completing the phone call, the participants were asked to complete an extensive health history and complete shipping and pickup information for the collected samples. Once the participants were fully enrolled, they were sent a study kit (see *Study Kit Components* section for details). Upon receiving their study kit, participants were instructed to collect their first blood and nasal samples as soon as possible.

### Study Design

The study was divided into three portions: (1) month 1 sampling, (2) extended sampling for select participants, and (3) follow-up symptom surveys and is summarized in **Figure 1**. During the first month of the study, participants collected a set of samples every 5 days. At each time point (1 - 5), participants collected and stabilized a blood sample (see *Blood Collection* section for details) and a nasal swab (see *Nasal Swab Collection* section for details). At time points and 1 and 5 (first and last samples of the month 1 sampling) participants also collected one additional unstabilized blood sample. Following sampling completion, participants filled out a survey about their COVID-19 symptoms and the sampling. The samples were then packaged and sent back to the lab (see *Sample Shipping* section for details) where they were kept at -20°C until further processing (see *Sample Processing* section for details). For more details about the extended sampling, see *Supplement 1, section*

**Figure 1.**
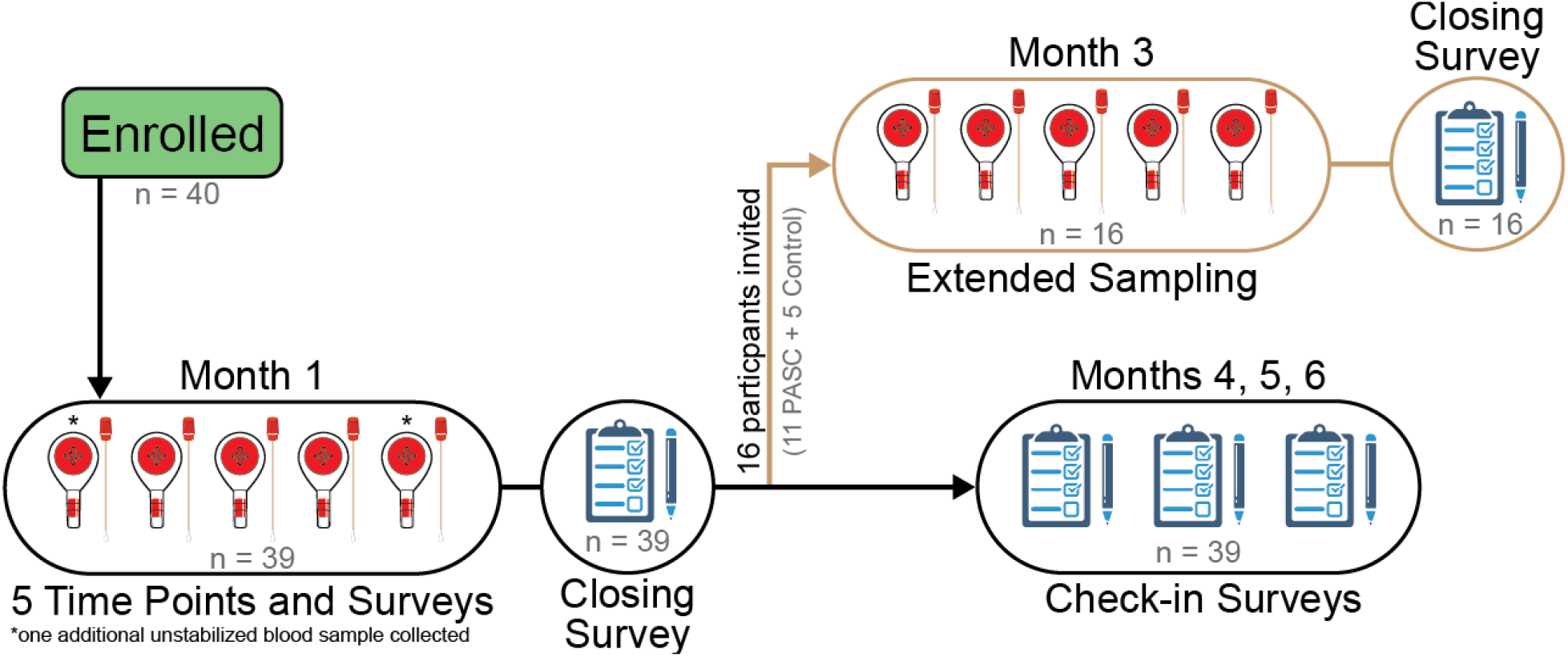
Outline of the study timeline. After enrollment, participants were sent a study kit with materials needed for completing the first five sample time points over the course of the first month of their infection. Over the course of the first month, participants collected a total of 7 blood samples and 5 nasal swabs. Participant symptoms were tracked and those who were mostly likely to develop PASC, along with age- and BMI-matched controls, were invited to the extended sampling which consisted of an additional 5 blood samples and 5 nasal swabs during month 3. Each time point was accompanied by a sampling survey with details about sampling time, blood volume collected, and experience using the kit. All participants were resurveyed for symptoms at months 4, 5, and 6. At time samples 5 and 10, participants were also given a closing survey with questions about overall study experience.

All participants (*n* = 39) were also surveyed for their overall wellbeing and symptoms at months 4, 5, and 6.

### Study Kit Components

The study kit included all the materials necessary for sampling and consisted of (1) individualized schedule for sampling and welcome letter, (2) copies of instructions for use (IFUs) for the blood collection and nasal swabs, (3) kits with sampling materials for time points 1 - 5 (or 6 - 10 for extended sampling), (4) 2 Tasso-SST kits (note: not included in kit for extended sampling), (5) ThermoPro TP49 temperature and humidity monitor, (6) backup materials, and (7) 5 pre-labeled UPS Labpaks. For a view of the fully prepared package, see **Figure e3**.

### Blood Collection and Stabilization

Blood collection and stabilization were performed using the standard homeRNA protocol.^10^ The additional blood samples at timepoints 1 and 5 that are not stabilized were collected according to the Tasso-SST manufacturer’s instructions. The collected samples were then placed in a 95kPa biohazard bag. The IFUs for the blood collection and stabilization can be found in **Supplement 2**.

### Sample Shipping

Once all the samples were collected and placed in the 95kPa biohazard bag, the biohazard bag was sealed and placed in the original time point box. The box containing the samples was then placed into a paid and pre-labeled UPS LabPak. The LabPak was left outside the following morning for courier pickup which was scheduled by the study team.

### Sampling Surveys

Following each sampling, participants completed sample surveys. Here, they were asked about their ongoing symptoms and questions about the sampling, including sampling time, blood collected, ease of use, etc. For the full survey, see **Supplement 2**.

### Closing Surveys

The closing surveys were administered both at sampling time point 5 and time point 10. The original version of the surveys at timepoint 5 was administered to the first 10 participants but was then reformulated using the Consolidated Framework for Implementation Research (CFIR), a determinants framework commonly used in healthcare settings to explore barriers and facilitators to the adoption of evidence-based interventions.^25^ Participants who filled out the original version were resurveyed with the updated closing survey within two months. To characterize the usability of the homeRNA platform in U3 populations, we focused on the Innovation Domain and put special emphasis on the Innovation Evidence-Base, Innovation Relative Advantage, and Innovation Design constructs. These were selected to explore the perceived effectiveness of homeRNA, the ease of use compared to in-person clinic visits, and the overall experience with the kit usage. The closing survey also includes questions from the Harvard Flourish Scale to assess the well-being of our participants.^26^

### Study Compensation

Study compensation was issued each time participants completed a survey. For time points that included sample collections, participants earned $20 each time, for a total of $100 for month 1 sampling and an additional $100 for extended sampling. Monthly check-in surveys were compensated at $10 each for a total of $30 per participant. The study compensation was issued in the form of digital Tango gift cards. Additionally, each study kit included a University of Washington branded mug as a souvenir for participating in the study.

### Survey Analysis

Sampling and closing surveys were used for the usability analysis of the study design and the homeRNA platform. The data was transferred from REDCap into R. The data manipulation was handled using the tydiverse library and data visualization was done with ggplot, GraphPad’s Prism, and SankeyMATIC.

## Results

In this study, we tracked participant symptoms for 6 months after the initial onset of symptoms, taking care to recruit participants as close to their first positive COVID-19 test as possible. In the process of recruiting, we received a total of 334 complete responses, of which 65 met all inclusion criteria to participate in our study and were invited to participate (**Figure 2A**). A total of 44 participants completed the consent process, of which 40 completed at least one sampling and were considered to be fully enrolled (for details on blood collection time and volume, see **Figure e4**). Most of these remaining participants (98%, *n* = 39/40) completed all sampling timepoints, surveys, and follow-ups, with one participant being withdrawn (lost to followup) from the study after completing collection of the first three samples. We were able to recruit participants from 19 states (**Figure 2B, Table 1**) with a diverse racial, ethnic, and age distribution (**Figure 2C, Table 1**), most of whom (80%, **Figure 1C**) are categorized as having a disadvantaged background per the NIH (see **Supplement 2** for NIH disadvantaged background criteria).

**Table 1.** Participant demographics.

| Characteristic | All<br>Participants<br>(n = 39) | Extended Study participants (n = 16) |  |
| --- | --- | --- | --- |
|  |  | PASC Predicted<br>(n = 11) | Non-PASC Control<br>(n = 5) |
| Sex<sup>1</sup> |  |  |  |
| Male | 0 | 0 | 0 |
| Female | 39 (100%) | 11 (100%) | 5 (100%) |
| Age (years)<sup>1</sup> | 43 (14) | 44 (13) | 58 (10) |
| Gender |  |  |  |
| Woman | 37 (95%) | 11 (100%) | 5 (100%) |
| Man | 0 (0%) | 0 (0%) | 0 (0%) |
| Transgender | 0 (0%) | 0 (0%) | 0 (0%) |
| Nonbinary | 2 (5%) | 0 (0%) | 0 (0%) |
| Race<sup>1</sup> |  |  |  |
| White, Middle Eastern or North African | 24 (62%) | 5 (45%) | 3 (60%) |
| American Indian or Alaskan Native | 2 (5%) | 1 (9%) | 1 (20%) |
| Black or African-American | 6 (15%) | 2 (18%) | 0 (0%) |
| Asian | 4 (10%) | 2 (18%) | 1 (0%) |
| More than one race | 1 (3%) | 1 (9%) | 2 (0%) |
| Other | 2 (5%) | 0 (0%) | 1 (20%) |
| Ethnicity<sup>1</sup> |  |  |  |
| Hispanic | 6 (15%) | 1 (9%) | 1 (20%) |
| Non-Hispanic | 33 (85%) | 10 (91%) | 4 (80%) |
| COVID-19 Vaccination Status<sup>1</sup> |  |  |  |
| Vaccinated | 34 (87%) | 9 (82%) | 4 (80%) |
| Not Vaccinated | 5 (13%) | 2 (18%) | 1 (20%) |
| Number of COVID Infections<br>(including present infection)<sup>2</sup> | 2 (0.86) | 2 (1) | 1 (0) |
| BMI<sup>2</sup> | 30 (7.2) | 30 (7.1) | 30 (4.8) |
| <sup>1</sup>n (%), <sup>2</sup>mean (SD) |  |  |  |

**Figure 2.**
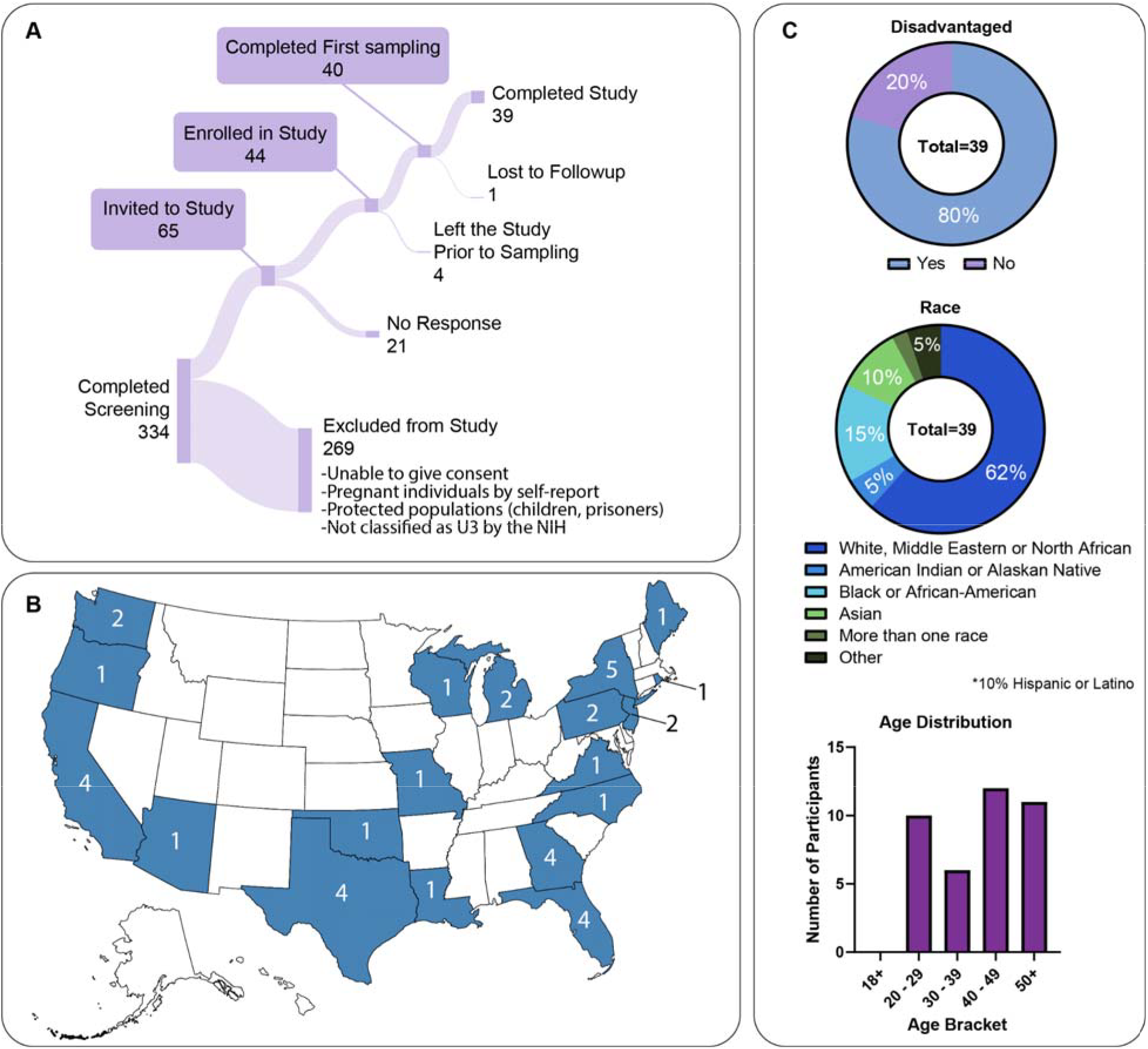
Outline of participant recruitment and demographics. (A) Sankey diagram depicting screening, enrollment, and retention in the study, (B) map of the United States with numbers of participants from each state, and (C) demographics of study participants including disadvantaged background per NIH, race, and age bracket.

Considering PASC predictive factors outlined in the *Methods* section, a subset of 11 individuals were predicted to be likely to develop PASC. These participants, along with a smaller subset of 5 controls who were age- and BMI-matched as closely as possible to the 11 PASC predicted participants, were invited to participate in an additional set of 5 sampling timepoints over the third month of their infection.

All participants who completed the original 5 timepoints, as well as those who participated in the extended sampling (*n* = 39 total) were surveyed at months 4, 5, and 6 from initial symptom onset and all 39 participants completed these follow-up surveys.

The closing survey at timepoint 5 included questions about the ease of use of the homeRNA platform, as well as general study experience. High participant satisfaction was illustrated (**Figure 3**) by the willingness of the cohort to participate in future research, with all (*n* = 39/39) participants indicating that they would be willing to participate in this same study or a similar study again, with 95% (*n* = 37/39) of participants agreeing or strongly agreeing that they enjoyed participating in this study. A high percentage (92%, *n* = 36/39) of participants either agreed or strongly agreed that they perceived participation in a remote study to be easier than a study that required in-person visits. Most participants (82%, *n* = 32/39) indicated they would be willing to participate for up to 4 years in a similar study with 15 samples collected per year.

**Figure 3.**
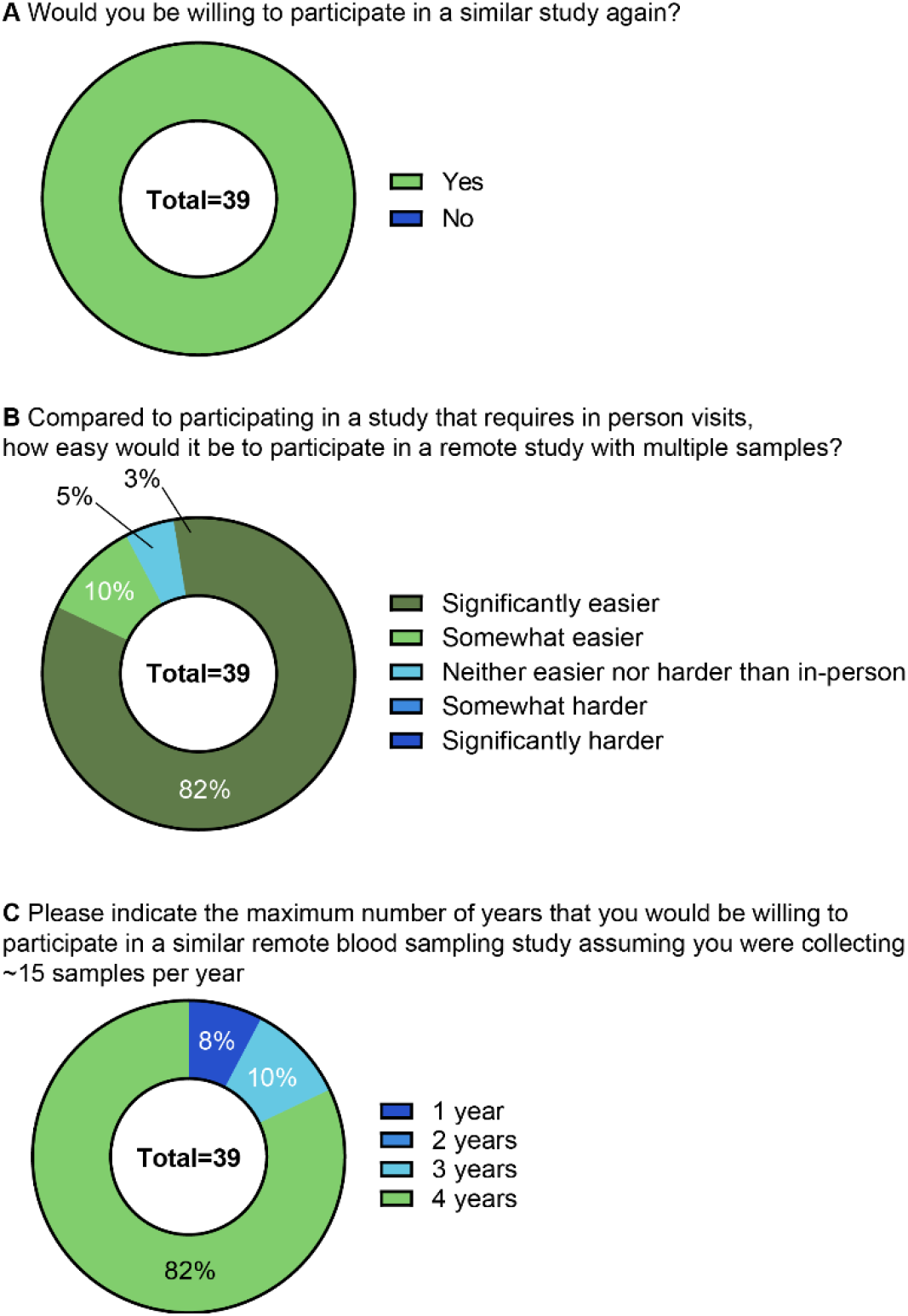
Summary of closing survey responses. All participants (*n* = 39/39) indicated that they would participate in a similar study again (A). Most participants (92%) indicated that a remote study would be easier or significantly easier than an in-person one. Most participants (82%) also indicated that they would be willing to participate in a remote study with 15 samples collected per year up to four years, which was the longest option available.

Beyond the flexibility of the remote design, study experience hinges on the sampling materials. Overwhelmingly, participants found the kit components easy to use, with the majority (>90%) indicating that they either agreed or strongly agreed with each of the statements outlined in **Figure 4**, with only one participant disagreeing for a single component (Tasso-SST instructions). The participants similarly reported enjoying participating in the study with 77% (*n* = 30/39) strongly agreeing and 18% agreeing (*n* = 7/39). Furthermore, our design included a phone call prior to enrollment to provide participants with an opportunity to connect with a study coordinator, ask questions about the study, and voice any concerns. A high percentage (82%, *n* = 32/39) reported that they strongly agreed that the study phone call was helpful, with an additional 10% (*n* = 4/49) indicating that they agreed.

**Figure 4.**
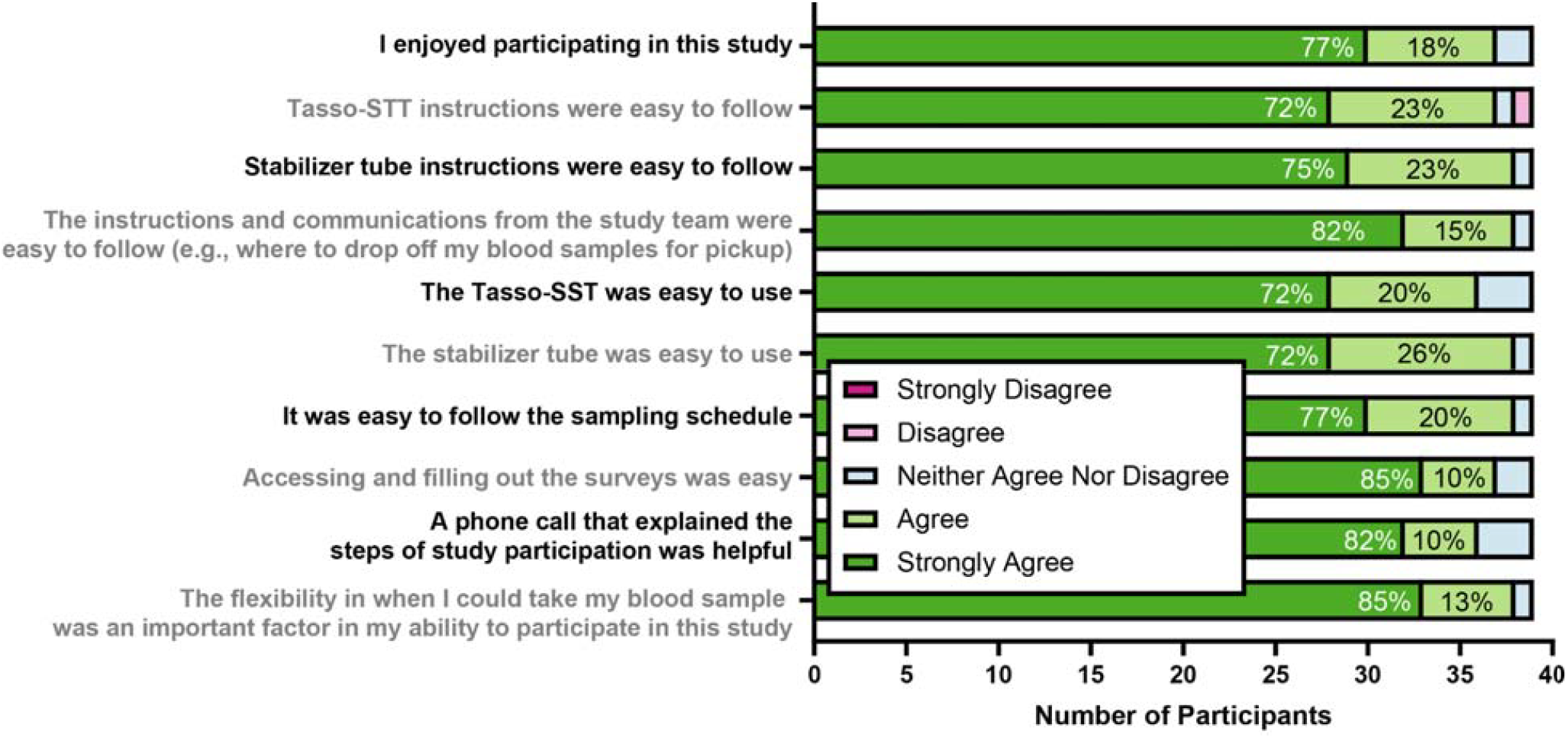
Summary of participant study experience. Each bar on the graph represents one question from the closing survey, with each of the sections corresponding to level of agreement for each statement. Overall, the majority of the participants (>90%) agreed or strongly agreed that they enjoyed participating in the study and that each of the components of the homeRNA kit was easy to use. Only one person reported finding the Tasso-SST difficult to use.

Sampling flexibility may be a strong component of the participants’ positive perception of the study. This was reflected in the high percentage (92%, *n* = 36/39) of participants who either agreed or strongly agreed that they perceived participation in a remote study to be easier than a study that required in-person visit (**Figure 3B**).

An additional consideration for study design includes participant motivation. Our participants were asked to indicate all factors they considered when joining the study. The strongest motivator in this cohort was the research topic, with 85% indicating an interest in adding to the body of knowledge on the health impacts of COVID-19 as a reason for joining the study, followed closely by flexibility of a remote study (69%). Participants were also asked about their motivation for continuing in the study. They similarly indicated an interest in adding to the body of knowledge on the health impacts of COVID-19 (87%), flexibility of a remote study (67%), and positive interactions with the study team (59%) as motivators for their continued participation. Total number of responses can be found in **Figure e5**. It is also worth noting that this work was conducted at a time when interest in COVID research was waning nationally and free COVID tests were being discontinued.

For participants who were selected for the extended sampling, another closing survey was administered at time point 10. The answers from these questions were analyzed using the same methods and can be found in **Figure e6**. The only notable changes were that at time point 10, no participants rated any of the kit components as difficult to use, and one person changed their answer for how long they would be willing to do a similar study from 3 years (survey at time point 5) to 4 years (survey at time point 10).

## Discussion

In recent years, there has been an effort to determine factors and design considerations for conducting more inclusive clinical studies.^3,5,27–30^ Some data have been collected on populations that qualify as U3 by the NIH definition and this has been an important step in balancing recruitment in clinical studies. Our work contributes to these efforts by conducting a longitudinal study with complex molecular readouts and surveying our U3 cohort for motivation, study experience, and willingness to participate in future studies. The existing literature and our past experience conducting remote studies indicate that logistical barriers such as transportation, commute length, and scheduling prevent participation in research. One approach to overcoming these barriers is to shift the sample collection from a centralized clinic, research institute, or blood collection site to the participants’ homes. The homeRNA platform, an at-home blood self-collection and RNA stabilization kit, enables this shift for longitudinal studies with transcriptomic readouts.

The successful implementation of the homeRNA platform in U3 populations is indicated by the high percentage (100%, *n* = 39/39) of participants who reported they would participate in a similar study again (**Figure 3A**). Participants also overwhelmingly reported that they enjoyed participating in the study and that homeRNA was easy to use (**Figure 4**). The sampling flexibility was an important factor in participants’ ability to be in the study (98%), and the initial phone call was helpful (92%). The sampling flexibility (time of day, location) overcame the reported barriers to in-person study participation (**Figure e7**), and the initial study phone call established rapport between the participants and study coordinators. We strongly suspect that these two were key components of the study and helped with retention and participant satisfaction. We note that we had a greater number of calls from participants at study launch than with later recruits, which we attribute to the addition of more detailed instructions in the sampling reminder emails. In other words, we found that clear and detailed instructions, even when redundant, helped the participants understand the study procedures. A full summary of materials provided to participants can be found in **Supplement 2**.

Furthermore, one of the more striking results of this work was the participant willingness to participate long-term in a remote clinical study (**Figure 3C**), with 82% (*n* = 32/39) indicating they would be willing to participate for up to 4 years in a similar study with 15 samples collected per year. This was the longest time period that participants were able to select in their surveys, suggesting that they may be willing to participate in even longer studies. In fact, in a follow-up study many participants from this cohort indicated that they would be willing to contribute to a study for “as long as necessary.” These survey data point to the success of the remote model in reaching U3 populations and suggest that remote studies can be valuable tools for longer-term longitudinal studies.

## Limitations

While we were able to gather comprehensive data about participant symptoms, predicting likelihood of long-term symptoms was a major challenge. At the time of the study, there was limited and evolving information about the definition of PASC and the symptoms associated with it. A limitation of the study presented here is that we have not yet analyzed the nasal swabs to confirm COVID-19 infection status and that we do not know if there were any concurrent respiratory infections that could be contributing to the reported symptoms. As these samples are analyzed, our data will become more useful in understanding symptom progression. Moreover, the quality of the sequencing data from blood RNA will rely heavily on how well the samples were stabilized by the participants. Sample processing is currently underway and the preliminary results show good RNA quality and yields.

In terms of study logistics, one limitation of our design was recruitment. Since we primarily advertised through social media, individuals who would otherwise be eligible but did not know about our study would not be recruited. In other studies that focus on reaching U3 populations, it may be useful to adopt a hybrid approach where advertising is done both online and with physical media including flyers, posters, and newspapers. Finally, it is important to note that while the remote, homeRNA-based study model overcomes logistical barriers to participation, more work needs to be done to overcome the larger cultural and systemic issues that have resulted in the underrepresentation of certain populations in clinical research.

## Conclusion

The homeRNA-based remote longitudinal study model was successful in a cohort of COVID-19+ U3 women. Survey responses indicated high satisfaction with study experience with excellent retention (98%, *n* = 39/40) over 6 months, despite the complexity and burden of the study. This success was attributed to a combination of the flexibility of remote sampling, communication with the study team, and ease of use of the sampling kit components. As such, we conclude that the remote study model outlined here can be a powerful tool for reaching participants from U3 populations who might otherwise be unlikely to participate in scientific or clinical studies. In ongoing work, we are further characterizing this cohort and their motivations through additional surveys and interviews to better understand the factors that contributed to the excellent participant retention and engagement. Ultimately, we hope that this body of work will serve as a roadmap for designing flexible and inclusive longitudinal studies with remote biospecimen collection.

## Supporting information

Supplement 1 - Figures and Definitions

Supplement 2 - Participant-Facing Materials

## Data Availability

The reported data are contained within the manuscript and supplementary files.

## References

1. Robiner WN, Yozwiak JA, Bearman DL, Strand TD, Strasburg KR. Barriers to clinical research participation in a diabetes randomized clinical trial. Social Science & Medicine. 2009;68(6):1069–1074. doi:10.1016/j.socscimed.2008.12.025

2. Marcantonio ER, Aneja J, Jones RN, et al. Maximizing Clinical Research Participation in Vulnerable Older Persons: Identification of Barriers and Motivators. Journal of the American Geriatrics Society. 2008;56(8):1522–1527. doi:10.1111/j.1532-5415.2008.01829.x

3. Luebbert R, Perez A. Barriers to Clinical Research Participation Among African Americans. J Transcult Nurs. 2016;27(5):456–463. doi:10.1177/1043659615575578

4. Hensen B, Mackworth-Young CRS, Simwinga M, et al. Remote data collection for public health research in a COVID-19 era: ethical implications, challenges and opportunities. Health Policy Plan. 2021;36(3):360–368. doi:10.1093/heapol/czaa158

5. Coakley M, Fadiran EO, Parrish LJ, Griffith RA, Weiss E, Carter C. Dialogues on diversifying clinical trials: successful strategies for engaging women and minorities in clinical trials. J Womens Health (Larchmt). 2012;21(7):713–716. doi:10.1089/jwh.2012.3733

6. Nomali M, Mehrdad N, Heidari ME, et al. Challenges and solutions in clinical research during the COVID-19 pandemic: A narrative review. Health Science Reports. 2023;6(8):e1482. doi:10.1002/hsr2.1482

7. Loucks TL, Tyson C, Dorr D, et al. Clinical research during the COVID-19 pandemic: The role of virtual visits and digital approaches. Journal of Clinical and Translational Science. 2021;5(1):e102. doi:10.1017/cts.2021.19

8. Chaussabel D, Pulendran B. A vision and a prescription for big data–enabled medicine. Nat Immunol. 2015;16(5):435–439. doi:10.1038/ni.3151

9. Rinchai D, Deola S, Zoppoli G, et al. High–temporal resolution profiling reveals distinct immune trajectories following the first and second doses of COVID-19 mRNA vaccines. Science Advances. 2022;8(45):eabp9961. doi:10.1126/sciadv.abp9961

10. Haack AJ, Lim FY, Kennedy DS, et al. homeRNA: A Self-Sampling Kit for the Collection of Peripheral Blood and Stabilization of RNA. Anal Chem. 2021;93(39):13196–13203. doi:10.1021/acs.analchem.1c02008

11. Stefanovic F, Brown LG, MacDonald J, et al. Your Blood is Out for Delivery: Considerations of Shipping Time and Temperature on Degradation of RNA from Stabilized Whole Blood. Anal Chem. 2025;97(3):1635–1644. doi:10.1021/acs.analchem.4c04591

12. Lim FY, Kim SY, Kulkarni KN, et al. High-frequency home self-collection of capillary blood correlates IFI27 expression kinetics with SARS-CoV-2 viral clearance. J Clin Invest. 2023;133(23). doi:10.1172/JCI173715

13. Haack AJ, Brown LG, Zeng Y, et al. A Flexible and Responsive Remote Study Design to Assess Gene Expression Changes During Wildfire Smoke Exposure with homeRNA, an At-home Blood Sampling Kit. medRxiv. Preprint posted online October 14, 2025:2025.10.11.25337783. doi:10.1101/2025.10.11.25337783

14. Lim FY, Lea HG, Dostie AM, et al. homeRNA self-blood collection enables high-frequency temporal profiling of presymptomatic host immune kinetics to respiratory viral infection: a prospective cohort study. eBioMedicine. 2025;112. doi:10.1016/j.ebiom.2024.105531

15. Brown LG, Haack AJ, Kennedy DS, et al. At-home blood collection and stabilization in high temperature climates using homeRNA. Front Digit Health. 2022;4:903153. doi:10.3389/fdgth.2022.903153

16. Sudre CH, Murray B, Varsavsky T, et al. Attributes and predictors of long COVID. Nat Med. 2021;27(4):626–631. doi:10.1038/s41591-021-01292-y

17. Raman B, Bluemke DA, Lüscher TF, Neubauer S. Long COVID: post-acute sequelae of COVID-19 with a cardiovascular focus. Eur Heart J. 2022;43(11):1157–1172. doi:10.1093/eurheartj/ehac031

18. Parotto M, Gyöngyösi M, Howe K, et al. Post-acute sequelae of COVID-19: understanding and addressing the burden of multisystem manifestations. The Lancet Respiratory Medicine. 2023;11(8):739–754. doi:10.1016/S2213-2600(23)00239-4

19. Al-Aly Z, Xie Y, Bowe B. High-dimensional characterization of post-acute sequelae of COVID-19. Nature. 2021;594(7862):259–264. doi:10.1038/s41586-021-03553-9

20. Thaweethai T, Donohue SE, Martin JN, et al. Long COVID trajectories in the prospectively followed RECOVER-Adult US cohort. Nat Commun. 2025;16(1):9557. doi:10.1038/s41467-025-65239-4

21. Wisk LE, L’Hommedieu M, Diaz Roldan K, et al. Variability in Long COVID Definitions and Validation of Published Prevalence Rates. JAMA Netw Open. 2025;8(8):e2526506. doi:10.1001/jamanetworkopen.2025.26506

22. Bowe B, Xie Y, Al-Aly Z. Postacute sequelae of COVID-19 at 2 years. Nat Med. 2023;29(9):2347–2357. doi:10.1038/s41591-023-02521-2

23. Cai M, Xie Y, Topol EJ, Al-Aly Z. Three-year outcomes of post-acute sequelae of COVID-19. Nat Med. 2024;30(6):1564–1573. doi:10.1038/s41591-024-02987-8

24. Pre-existing conditions associated with post-acute sequelae of COVID-19 | Elsevier Enhanced Reader. doi:10.1016/j.jaut.2022.102991

25. Damschroder LJ, Reardon CM, Widerquist MAO, Lowery J. The updated Consolidated Framework for Implementation Research based on user feedback. Implementation Science. 2022;17(1):75. doi:10.1186/s13012-022-01245-0

26. Our Flourishing Measure | The Human Flourishing Program. Accessed October 17, 2025. https://hfh.fas.harvard.edu/measuring-flourishing

27. Wendler D, Kington R, Madans J, et al. Are Racial and Ethnic Minorities Less Willing to Participate in Health Research? PLOS Medicine. 2005;3(2):e19. doi:10.1371/journal.pmed.0030019

28. Girardi G, Longo M, Bremer AA. Social determinants of health in pregnant individuals from underrepresented, understudied, and underreported populations in the United States. Int J Equity Health. 2023;22(1):186. doi:10.1186/s12939-023-01963-x

29. Stewart J, Krows ML, Schaafsma TT, et al. Comparison of Racial, Ethnic, and Geographic Location Diversity of Participants Enrolled in Clinic-Based vs 2 Remote COVID-19 Clinical Trials. JAMA Netw Open. 2022;5(2):e2148325. doi:10.1001/jamanetworkopen.2021.48325

30. Garber M, and Arnold RM. Promoting the Participation of Minorities in Research. The American Journal of Bioethics. 2006;6(3):W14–W20. doi:10.1080/15265160600686331

