## Supplement 1 - Figures and Definitions for "Research In Your Mailbox: Remote Blood Sampling Enables Longitudinal Studies in Underserved Groups"

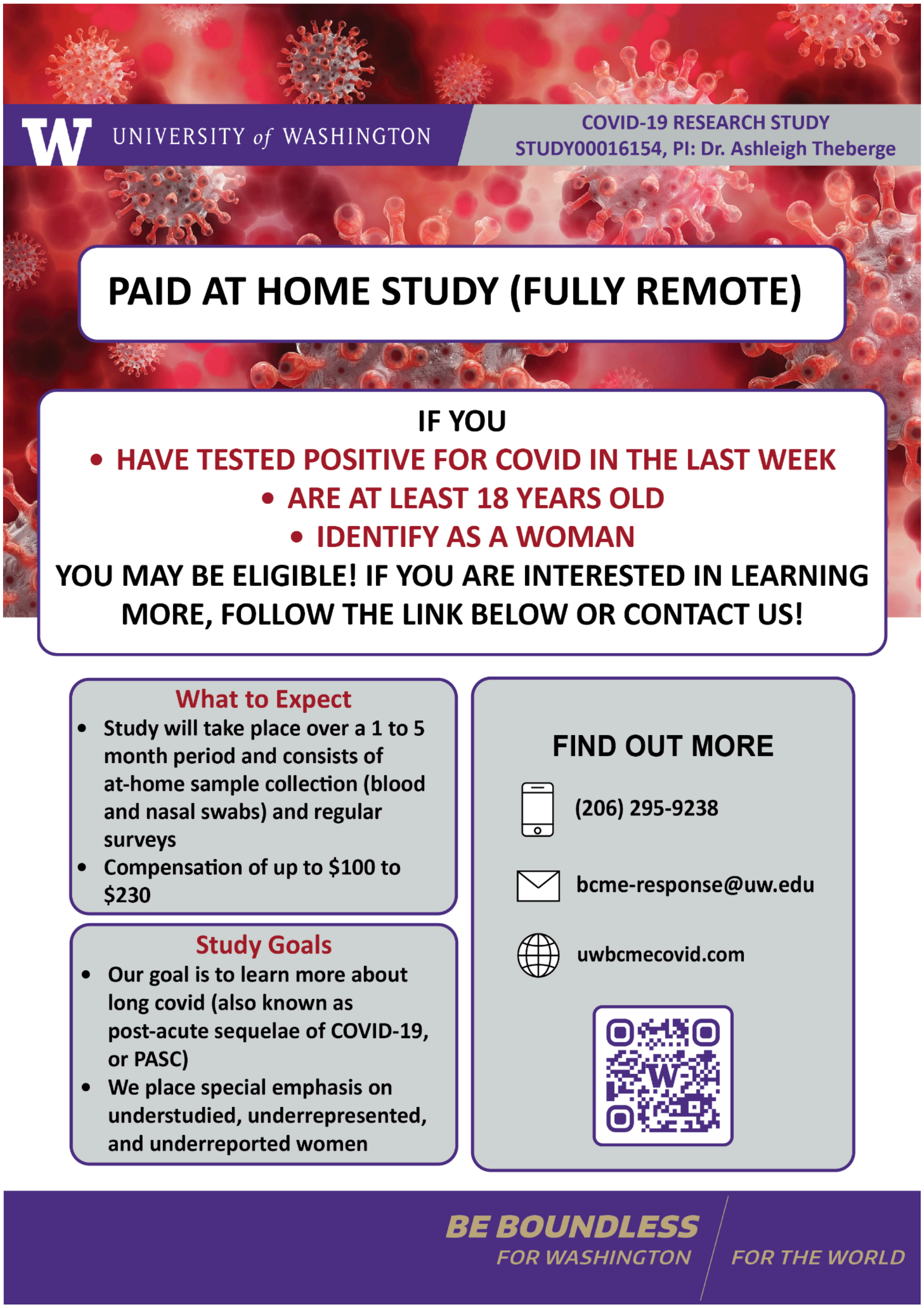


**Figure e1.** Example of a digital advertisement used for recruitment. Components that we have found important for our studies include time commitment, compensation, and location. Additional information such as inclusion criteria and study goals are useful additions provided that there is sufficient space.

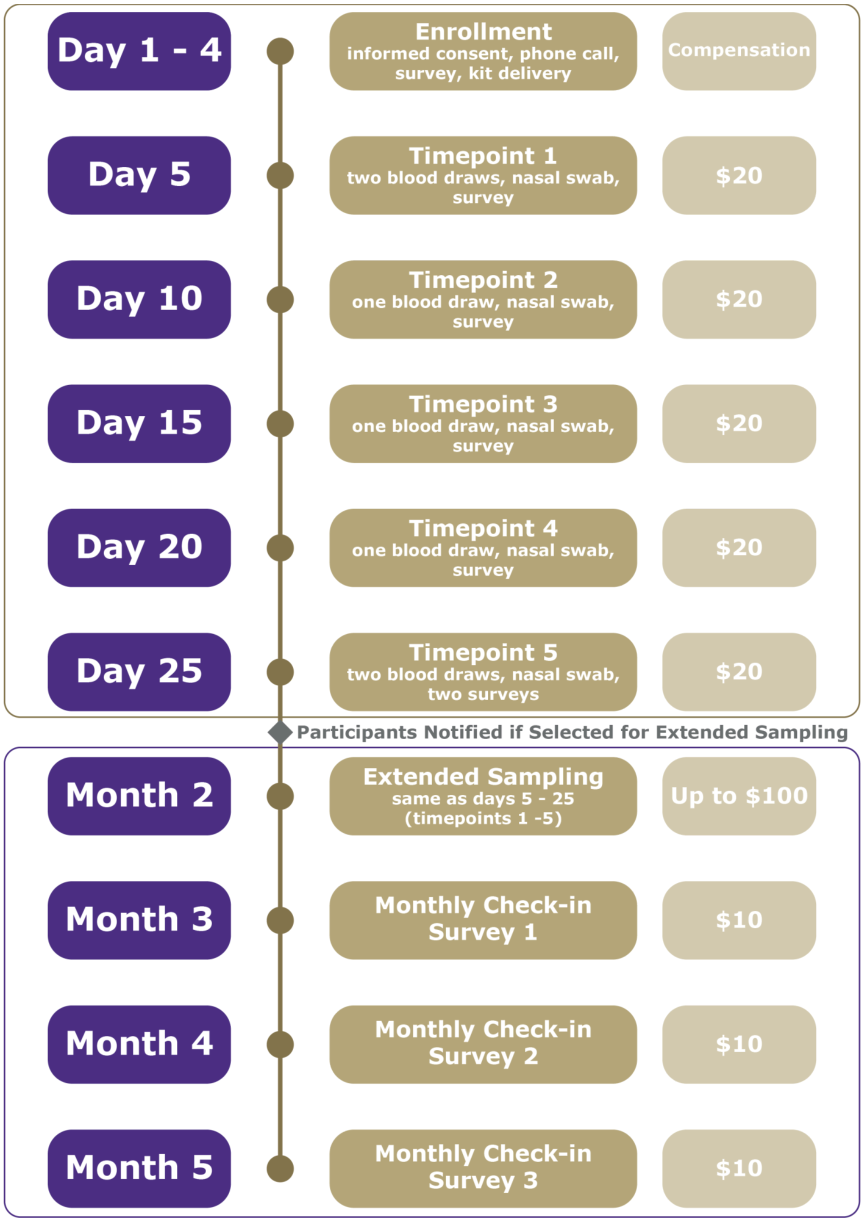


**Figure e2**. Study timeline provided to participants including breakdown of responsibilities at each time point and corresponding compensation.

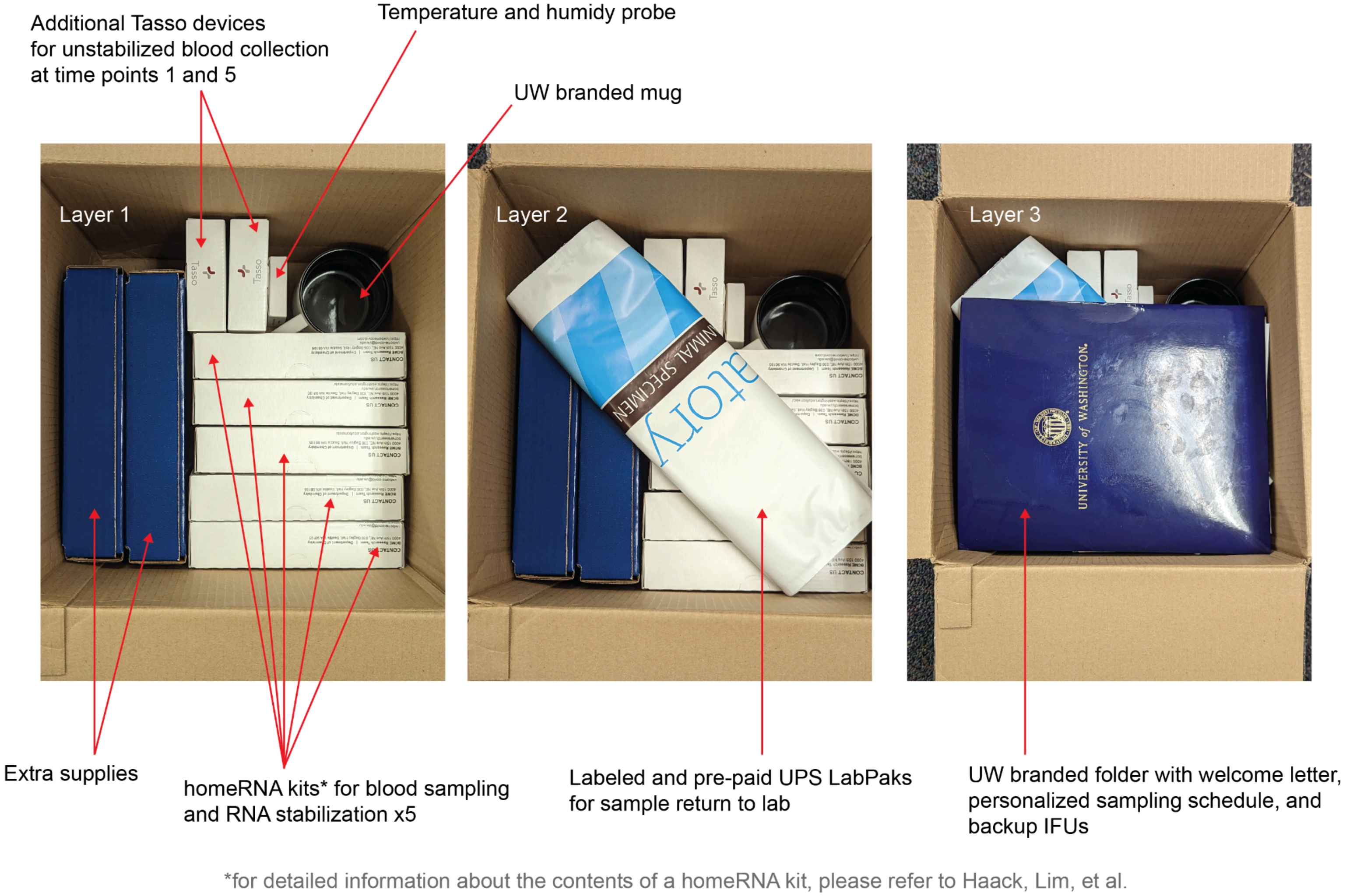


**Figure e3.** Study kit components. After enrollment in the study, participants were sent kits including all necessary study materials. This included 5 homeRNA kits (see Haack, Lim, et al. for details), 2 additional Tasso-SST devices for sampling without stabilization at time point 1 and 5, a temperature and humidity probe, a souvenir mug, 5 prepaid and labeled UPS LabPaks for sample return, and a folder including a welcome letter, personalized sampling timeline, and extra copies of instructions for use for each of the components. Each time point sampling box included materials needed to complete the blood draw, stabilization, and nasal swab collection. For blood draw and stabilization this included a Tasso-SST, homeRNA stabilizer tube filled with RNA*later*, Medline Instant Hot Pack Medium, a self-adhesive bandage, alcohol swab, and a 50 mL Falcon conical tube. The nasal swab collection components included one Sterile PurFlock Ultra Flocked Swab, Regular Tip, 6 in. Polystyrene Shaft and one Copan Diagnostics Universal Transport Medium (UTM-RT™) in Screw-Cap Tube. The backup materials included 2 additional duplicates of each component needed for sampling in case of issues with the original materials.

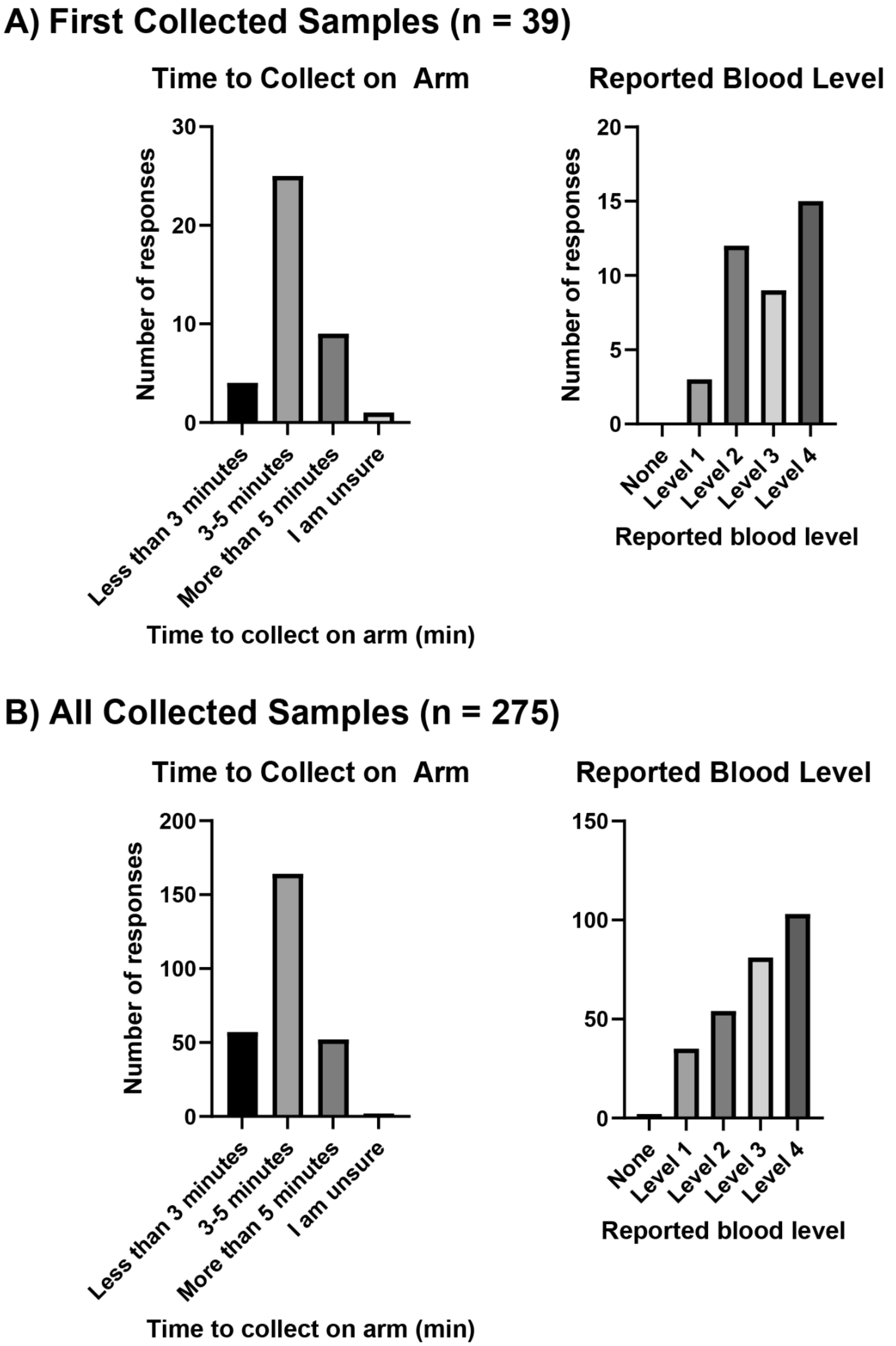


**Figure e4**. Summary of blood collection time and reported blood volume at first use (A) and across all samples (B). Overall, participants tended to spend roughly the same amount of time on blood collection throughout the study with fairly consistent blood volumes collected.

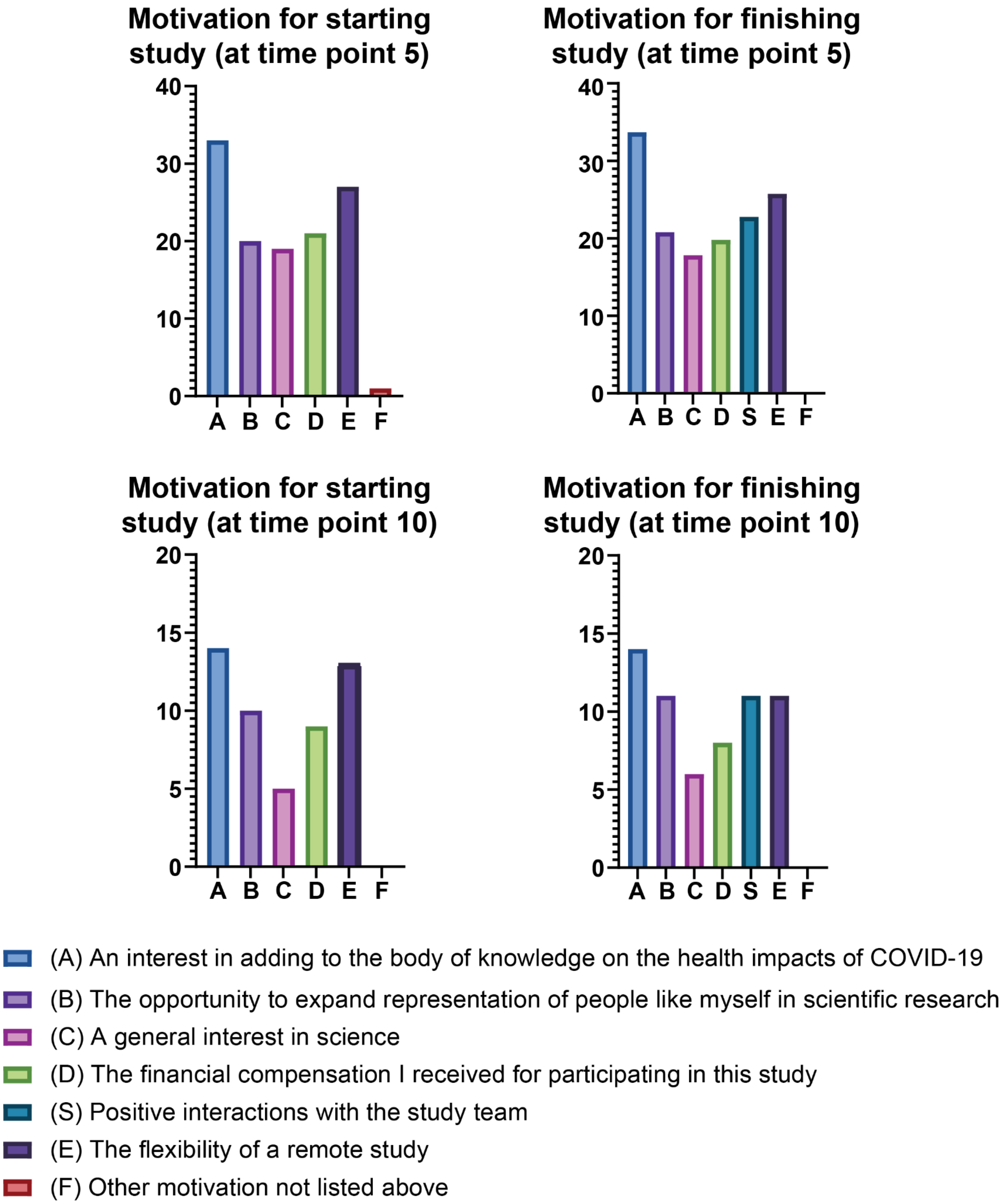


**Figure e5**. Summary of motivation for starting and continuing in study. The most important factors for starting in the study included interest in the topic (COVID-19) and flexibility of the remote study. In addition to these factors, participants reported positive interactions with the study team were a motivator for continuing in the study once enrolled. The answers of the whole cohort ($n$ = 39) at time point 5 and the extended sampling participants at time point 10 did not differ meaningfully.

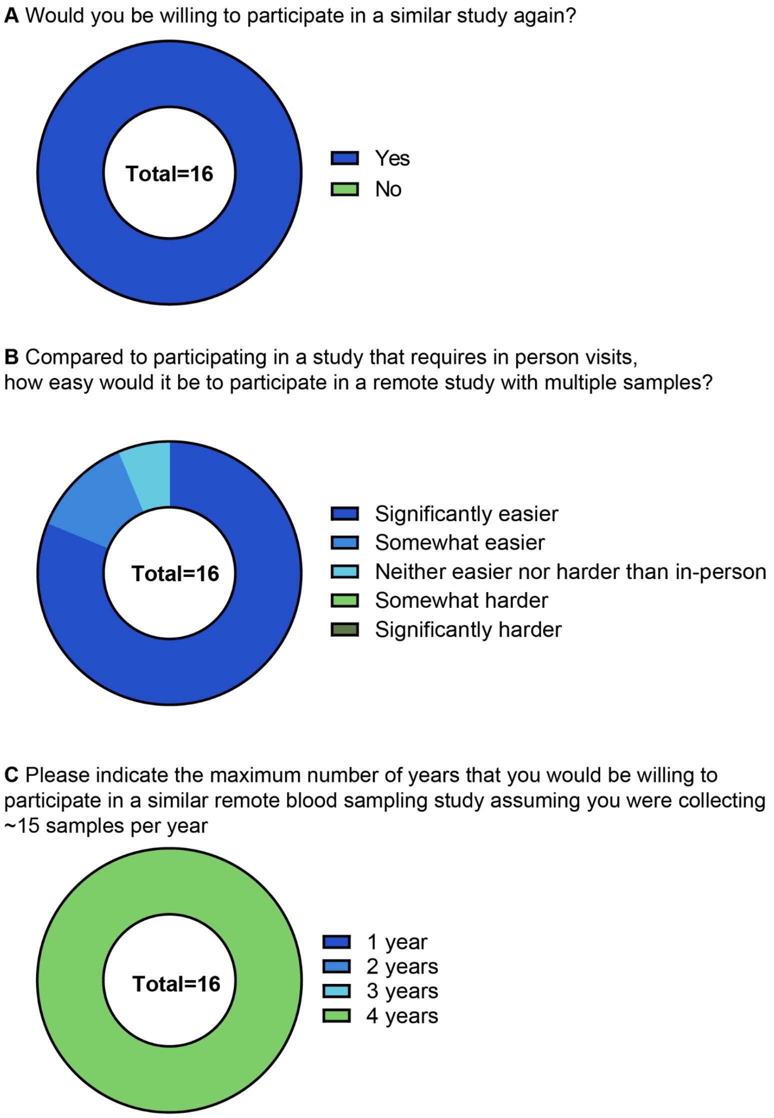


**Figure e6**. Summary of closing survey responses for extended sampling. All participants (*n* = 16/16) indicated that they would participate in a similar study again (A). Most participants (94%, *n* = 15/16) indicated that a remote study would be easier or significantly easier than an in-person one. All participants (*n* = 16/16) also indicated that they would be willing to participate in a remote study with 15 samples collected per year up to four years, which was the longest option available. Importantly, none of the extended study participants changed their answers negatively from their original responses to the closing survey at time point 5.

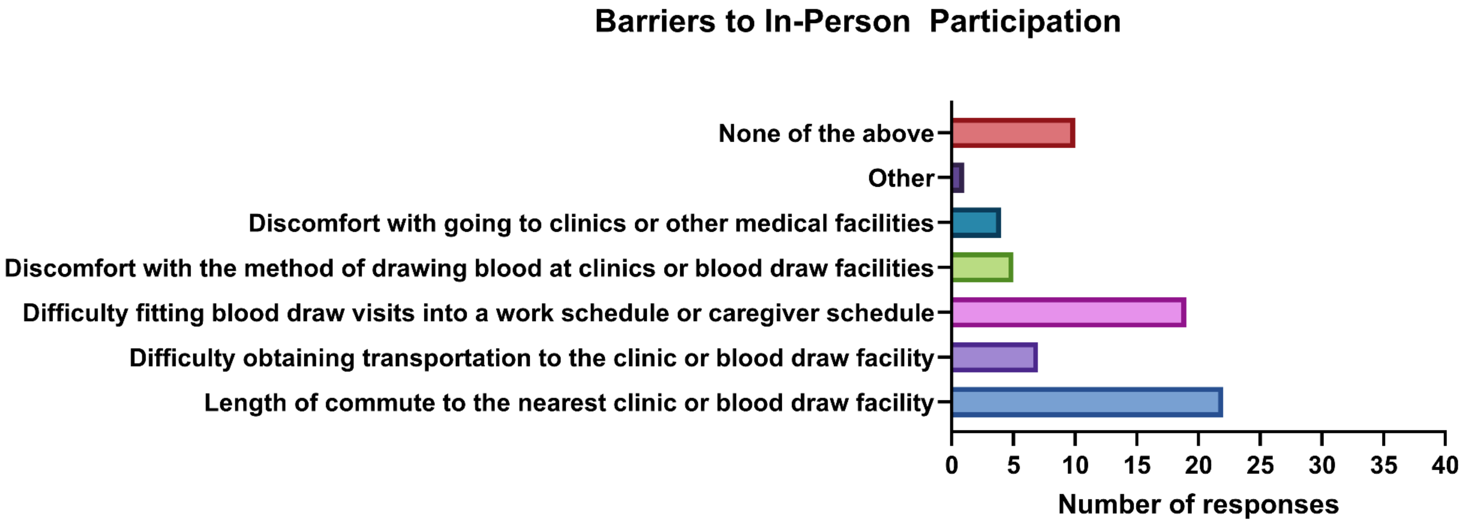


**Figure e7**. Summary of barriers to in-person participation.

**NIH Disadvantaged Background**
The following text was copied from the NIH eRA commons on 9/26/2025
(https://www.era.nih.gov/commons/disadvantaged_def.htm)

Disadvantaged Background
An individual is considered to be from a disadvantaged background if he or she meets two or more of the following criteria:

- Were or currently are homeless, as defined by the McKinney-Vento Homeless Assistance Act (Definition: https://nche.ed.gov/mckinney-vento/);
- Were or currently are in the foster care system, as defined by the Administration for Children and Families (Definition: https://www.acf.hhs.gov/cb/focus-areas/foster-care);
- Were eligible for the Federal Free and Reduced Lunch Program for two or more years (Definition: https://www.fns.usda.gov/school-meals/income-eligibility-guidelines);
- Have/had no parents or legal guardians who completed a bachelor's degree (see https://nces.ed.gov/pubs2018/2018009.pdf);
- Were or currently are eligible for Federal Pell grants (Definition: https://studentaid.gov/understand-aid/types/grants/pell);
- Received support from the Special Supplemental Nutrition Program for Women, Infants and Children (WIC) as a parent or child (Definition: https://www.fns.usda.gov/wic/wic-eligibility-requirements).
- Grew up in one of the following areas: a) a U.S. rural area, as designated by the Health Resources and Services Administration (HRSA) Rural Health Grants Eligibility Analyzer (https://data.hrsa.gov/tools/rural-health), or b) a Centers for Medicare and Medicaid Services-designated Low-Income and Health Professional Shortage Areas (qualifying zipcodes are included in the file). Only one of the two possibilities in #7 can be used as a criterion for the disadvantaged background definition.

**Supplemental Methods**

*Extended Sampling*

For the extended sampling, a total of 15 participants were chosen due to resource limitations. We opted to select a greater number of PASC-predicted individuals to maximize likelihood of capturing PASC, since it is possible for PASC-predicted participants to recover fully. Due to the longitudinal design of our study, data from PASC+ participants can be informative even in the absence of corresponding controls. Longitudinal sampling allows us to follow gene expression within a participant over time, rather than focusing on comparison between cases and controls at a single time point. This will be explored in further detail in subsequent publications examining the gene expression data resulting from this work.

Following the month 1 sampling, participants were assessed for likelihood for developing PASC based on their self-reported medical history and persistence of COVID-19 symptoms. This assessment was based on the PASC literature available at the time, where body mass index (BMI), pre-existing conditions (asthma, constipation, reflux, rheumatoid arthritis, seasonal allergies, and depression/anxiety), persistence of symptoms at one month (fatigue, headache, difficulty breathing, alterations in taste/smell, hoarse voice, muscle pain, and malaise), and more than 5 symptoms at time of onset made individuals more likely to develop PASC.[16,19,22,24] Participants who had more than 2 lingering symptoms, more than 5 reported symptoms at onset, and had at least one pre-existing condition or a BMI greater than 26 were considered for the PASC-predicted cohort. From this pool, participants were examined on a case-by-case basis, and 11 were selected as most likely to develop PASC. Any participant with ambiguities or low probability of developing PASC (e.g., a participant with only one pre-existing condition, healthy BMI, and who was vaccinated for COVID-19) were not selected. The pool of potential control participants was made by filtering for participants who met at least one of the following: no lingering symptoms, fewer than 5 symptoms at onset, no pre-existing conditions considered significant for PASC, or a high number of vaccine boosters (2+). To enable close correspondence between the control and PASC-predicted participants, control participants were enrolled based on roughly matching at least one of the PASC-predicted participants in age and BMI. A total of 4 controls were selected. The extended sampling ($n$ = 15) was set to start roughly 2 weeks after a participant's 5th sample and consisted of an additional 5 sampling time points (6 - 10) spaced 5 days apart with stabilized blood draws and nasal swabs (but not the additional, unstabilized blood draws). After each sampling, the participants completed sample surveys, and after time point 10 they completed an additional closing survey.

*Nasal Swabs*

Nasal swabs were collected by swirling the swab inside each nostril for 15 seconds. The swabs were then placed in the UTM-RT and snapped off. The UTM-RT tubes were then secured with the screw-top cap and added to the same 95kPa biohazard bag. The IFU for the nasal swab collection can be found in **Supplement 2**.

*Sample Processing*

When ready for sample processing, the samples were taken out of the -20°C freezers and transferred into the biosafety cabinet (BSC). The samples were carefully removed and the outside containers were sterilized first with ethanol then with trifectant spray. The stabilized blood samples and nasal swab samples were inventoried and transferred to -80°C until future processing. The unstabilized blood samples were centrifuged for 10 minutes at 15,000 RCF. Roughly 50 µL of the supernatant was removed and transferred into Screw Cap Micro Tubes (RNAse/DNAse free) and stored at -80°C for future analysis.
