## Supplement 2 - Participant-Facing Materials for "Research In Your Mailbox: Remote Blood Sampling Enables Longitudinal Studies in Underserved Groups"

Supplement 2  contains all digital and physical information shared with participants. Here is a sample timeline of when these would be seen by the participant:

-Prospective participant fills out Eligibility Survey and Screening Survey
-After survey review, prospective participants are sent an email invitation to consent
-Participants fill out the Informed Consent survey
-Participants are sent welcome survey invitation
-Participants fill out Welcome and Mailing Address surveys
-Participants are sent study kits and receive the welcome letter, sampling schedule, IFUs, and box inserts
-Once kit is received, participant is sent a sampling email invitation
-After sample collection, participants fill out sampling survey
-Future samplings use corresponding sampling email invitations and sampling surveys
-At time point 5, all participants also complete a closing survey
-Participants were evaluated for extended sampling
-Participants selected for extended sampling were sent another study kit with IFUs and extended letter
-At each extended sampling time point, participants received extended sampling email instructions
-After each extended sampling, participants completed extended sampling surveys
-At time point 10, participants who were selected for extended sampling completed an extended sampling closing survey
-At months 4, 5, and 6, all participants were sent an invitation for monthly check-in surveys and completed the corresponding surveys

*Original closing survey was updated and only the new version was administered to all participants. Participants who completed the original version were resurveyed with the updated closing survey.

### Eligibility Screen

Thank you for your interest in our study.

Please complete the survey below to determine if you qualify to participate in our study. We ask for your name and contact information for study enrollment purposes only. Should you decide not to participate, we will not retain your information.

---

What is your name?

First name: ______ Last name: ______

---

| | |
|---|---|
| Are you age 18 or older? | ◯ Yes<br>◯ No |
| What is your residential zip code? | ______________________ |
| What sex were you assigned at birth? | ◯ Male<br>◯ Female<br>◯ Intersex |
| What terms best express how you describe your gender identity? | ◯ Man<br>◯ Woman<br>◯ Transgender<br>◯ Non-binary<br>◯ None of these describe me, and I'd like to consider additional options |
| Are any of these a closer description to your gender identity? | ◯ Trans man/Transgender Man/FTM<br>◯ Trans woman/Transgender Woman/MTF<br>◯ Genderqueer<br>◯ Genderfluid<br>◯ Gender variant<br>◯ Questioning or unsure of your gender identity<br>◯ None of these describe me, and I want to specify<br>____________ |
| Please specify your gender identity: | ______________________ |
| Have you received a positive COVID-19 test result in the past week? | ◯ Yes<br>◯ No |
| What date were you tested? | ______________________ |
| If you have confirmation of the positive result, please upload it here. This can be a photo of an at home kit or documentation from a PCR test. | |
| Do you remember the date your symptoms started? | ◯ Yes<br>◯ No |
| What date did your symptoms start? | ______________________ |

Do any of these apply to you?
- I am pregnant
- I currently reside in a correctional facility
*Please inform the study team if this changes during your participation in the study

◯ Yes
◯ No

Thank you for your interest. Unfortunately, you do not qualify for the study at this time.

Please read the following description of the study protocol and expectations carefully:

This study will consist of a series of surveys and self-collected blood and nasal samples. The surveys will collect data about your overall health and your background, as well as information about sample collection. The blood sampling will be done using a Tasso-SSTTM device. The Tasso-SSTTM adheres to your upper arm and begins blood collection with the press of a button. The time needed for each blood collection is around 10 minutes. The nasal samples will be collected using a nasal swab like most PCR or at-home COVID-19 tests and should take around 5 minutes to complete. These samples will be taken at set timepoints and shipped back to the study team within 24-h of collection. At each sampling timepoint, you will collect up to two blood samples and one nasal swab. Most participants will be asked to complete 5 sampling timepoints, but a subset may be asked to participate in extended sampling for an additional 5 sampling timepoints and 3 additional surveys. You will be compensated in the form of digital gift cards based on sample collection and survey completion.

Are you still interested in participating?

◯ Yes ◯ No

What is your preferred name?

__________________________________
(Note: This is the name that will be used in any future correspondence relating to this study)

What is an e-mail address to receive study survey invitations, notifications and reminders?

__________________________________

What is a phone number the study team can reach you at?

__________________________________

Can this phone number can receive text messages (SMS)? ◯ Yes ◯ No

In general, when would be a good time for a one-time phone call to talk about the study details?

☐ Morning
☐ Midday
☐ Evening
(After signing the informed consent (ICF), please expect a phone call from our team within 48 hours. The number calling will be (206) 295-9238.)

How do you prefer to receive study survey invitations, notifications and reminders?

◯ Email
◯ Text
(Please note the phone number that sends the texts with study survey links and reminders is not a number we can be reached at. To contact the study team, please or text/call (206) 593-3073.)

Thank you for your time.

### U3/PASC Screen

| | |
|---|---|
| What is your age? | ________________ |
| What is your race or origin? (select all that apply) | ☐ American Indian or Alaskan Native<br>☐ Asian<br>☐ Black or African-American<br>☐ Native Hawaiian or Other Pacific Islander<br>☐ White, Middle Eastern or North African<br>☐ Other<br>☐ Prefer not to answer |
| Please specify | ________________ |
| Are you of Hispanic, Latino, or Spanish origin? (select all that apply) | ☐ No, not Hispanic, Spanish, Latino, or Spanish origin<br>☐ Yes, Mexican, Mexican-American, Chicano<br>☐ Yes, Puerto Rican<br>☐ Yes, other Hispanic, Latino, or Spanish origin<br>☐ Prefer not to answer |
| Which of the following best represents how you think of yourself at this time? | ☐ Straight<br>☐ Gay<br>☐ Lesbian<br>☐ Bisexual<br>☐ Other<br>☐ Prefer not to say |
| Have you been vaccinated for COVID-19? | ◯ Yes<br>◯ No |
| What was the brand(s) of the vaccine(s)? | ☐ Pfizer<br>☐ Moderna<br>☐ Johnson & Johnson<br>(check all that apply) |
| What are the approximate dates of your vaccinations? (Write in the form of MM-YY with each on a new line) | ________________ |
| Including the current infection, how many times have you had COVID-19? | ◯ 1<br>◯ 2<br>◯ 3<br>◯ 4<br>◯ 5<br>◯ 5+ |
| Not including the current infection, have you had COVID-19 in the past year? | ◯ Yes<br>◯ No |
| When (month and year) did your previous COVID-19 infection(s) start? | ________________ |
| Were you or are you currently homeless? | ◯ No<br>◯ Yes<br>◯ Not sure<br>◯ Prefer not to say |

| | |
|---|---|
| Were you ever in the foster care system? | ◯ No<br>◯ Yes<br>◯ Not sure<br>◯ Prefer not to say |
| Were you ever eligible for the Federal Free and Reduced Lunch Program for two or more years? | ◯ No<br>◯ Yes<br>◯ Not sure<br>◯ Prefer not to say |
| Have any of your parents or legal guardians completed a bachelor's degree? | ◯ No<br>◯ Yes<br>◯ Not sure<br>◯ Prefer not to answer |
| Were you ever or are you currently eligible for Federal Pell grants? | ◯ No<br>◯ Yes<br>◯ Not sure<br>◯ Prefer not to say |
| Have you ever received support from the Special Supplemental Nutrition Program for Women, Infants and Children as a parent or child? | ◯ No<br>◯ Yes<br>◯ Not sure<br>◯ Prefer not to say |

We will now show you a set of symptoms and will ask you in three different sections about their occurance, frequency, and severity. There may be instances where more than one answer could reflect your experience, in these cases, please select all that apply.

#### OCCURENCE OF SYMPTOMS (select all that apply)

| | I experienced this symptom as a result of my current COVID-19 infection | I experienced this symptom as a result of a prior COVID-19 infection | I experienced this symptom prior to any COVID-19 infection | I have not experienced this symptom | Not sure |
|---|---|---|---|---|---|
| Fatigue (Lack of energy or general tired feeling) | ☐ | ☐ | ☐ | ☐ | ☐ |
| Feeling tired after exercise (Symptoms worse after even minor physical or mental effort) | ☐ | ☐ | ☐ | ☐ | ☐ |
| Weakness in arms or legs | ☐ | ☐ | ☐ | ☐ | ☐ |
| Shortness of breath or difficult/labored breathing | ☐ | ☐ | ☐ | ☐ | ☐ |
| Feeling faint or dizzy; difficulty thinking soon after standing up from a sitting or lying position | ☐ | ☐ | ☐ | ☐ | ☐ |
| Chronic cough | ☐ | ☐ | ☐ | ☐ | ☐ |

| | | | | | |
|---|---|---|---|---|---|
| Congestion, sinus pain, sputum secretion | ☐ | ☐ | ☐ | ☐ | ☐ |
| Palpitations, racing heart, arrhythmia, skipped beats | ☐ | ☐ | ☐ | ☐ | ☐ |
| Tightness or pressure in chest | ☐ | ☐ | ☐ | ☐ | ☐ |
| Loss of or change in smell or taste | ☐ | ☐ | ☐ | ☐ | ☐ |
| Lack of appetite like you just haven't been hungry | ☐ | ☐ | ☐ | ☐ | ☐ |
| Abdominal pain | ☐ | ☐ | ☐ | ☐ | ☐ |
| Nausea or vomiting | ☐ | ☐ | ☐ | ☐ | ☐ |
| Diarrhea | ☐ | ☐ | ☐ | ☐ | ☐ |
| Constipation | ☐ | ☐ | ☐ | ☐ | ☐ |
| Reflux/GERD/heartburn | ☐ | ☐ | ☐ | ☐ | ☐ |
| Joint pain | ☐ | ☐ | ☐ | ☐ | ☐ |
| Muscle pain | ☐ | ☐ | ☐ | ☐ | ☐ |
| Headaches | ☐ | ☐ | ☐ | ☐ | ☐ |
| Problems thinking or concentrating ("brain fog") | ☐ | ☐ | ☐ | ☐ | ☐ |
| Nerve problems (tremor, shaking, abnormal movements, numbness, tingling, burning, can't move part of body, new seizures) | ☐ | ☐ | ☐ | ☐ | ☐ |
| Problems with anxiety, depression, stress, or trauma-related symptoms like nightmares or grief | ☐ | ☐ | ☐ | ☐ | ☐ |
| Problems with sleep | ☐ | ☐ | ☐ | ☐ | ☐ |

#### FREQUENCY OF SYMPTOMS
#### (PLEASE COMPLETE FOR PAST INFECTION(S) OR PRIOR TO ANY COVID INFECTION ONLY)

| | A few times a month or less | Once a week | A few times a week | Daily | Always or almost always | Not sure |
|---|---|---|---|---|---|---|
| Fatigue (Lack of energy or general tired feeling) | ○ | ○ | ○ | ○ | ○ | ○ |
| Feeling tired after exercise (Symptoms worse after even minor physical or mental effort) | ○ | ○ | ○ | ○ | ○ | ○ |
| Weakness in arms or legs | ○ | ○ | ○ | ○ | ○ | ○ |
| Shortness of breath or difficult/labored breathing | ○ | ○ | ○ | ○ | ○ | ○ |

| | | | | | | |
|---|---|---|---|---|---|---|
| Feeling faint or dizzy; difficulty thinking soon after standing up from a sitting or lying position | ○ | ○ | ○ | ○ | ○ | ○ |
| Chronic cough | ○ | ○ | ○ | ○ | ○ | ○ |
| Congestion, sinus pain, sputum secretion | ○ | ○ | ○ | ○ | ○ | ○ |
| Palpitations, racing heart, arrhythmia, skipped beats | ○ | ○ | ○ | ○ | ○ | ○ |
| Tightness or pressure in chest | ○ | ○ | ○ | ○ | ○ | ○ |
| Loss of or change in smell or taste | ○ | ○ | ○ | ○ | ○ | ○ |
| Lack of appetite like you just haven't been hungry | ○ | ○ | ○ | ○ | ○ | ○ |
| Abdominal pain | ○ | ○ | ○ | ○ | ○ | ○ |
| Nausea or vomiting | ○ | ○ | ○ | ○ | ○ | ○ |
| Diarrhea | ○ | ○ | ○ | ○ | ○ | ○ |
| Constipation | ○ | ○ | ○ | ○ | ○ | ○ |
| Reflux/GERD/heartburn | ○ | ○ | ○ | ○ | ○ | ○ |
| Joint pain | ○ | ○ | ○ | ○ | ○ | ○ |
| Muscle pain | ○ | ○ | ○ | ○ | ○ | ○ |
| Headaches | ○ | ○ | ○ | ○ | ○ | ○ |
| Problems thinking or concentrating ("brain fog") | ○ | ○ | ○ | ○ | ○ | ○ |
| Nerve problems (tremor, shaking, abnormal movements, numbness, tingling, burning, can't move part of body, new seizures) | ○ | ○ | ○ | ○ | ○ | ○ |
| Problems with anxiety, depression, stress, or trauma-related symptoms like nightmares or grief | ○ | ○ | ○ | ○ | ○ | ○ |
| Problems with sleep | ○ | ○ | ○ | ○ | ○ | ○ |

#### SEVERITY OF SYMPTOMS OF CURRENT INFECTION

| | Mild | Moderate | Severe |
|---|---|---|---|
| Fatigue (Lack of energy or general tired feeling) | ○ | ○ | ○ |
| Feeling tired after exercise (Symptoms worse after even minor physical or mental effort) | ○ | ○ | ○ |
| Weakness in arms or legs | ○ | ○ | ○ |
| Shortness of breath or difficult/labored breathing | ○ | ○ | ○ |

| | | | |
|---|---|---|---|
| Feeling faint or dizzy; difficulty thinking soon after standing up from a sitting or lying position | ○ | ○ | ○ |
| Chronic cough | ○ | ○ | ○ |
| Congestion, sinus pain, sputum secretion | ○ | ○ | ○ |
| Palpitations, racing heart, arrhythmia, skipped beats | ○ | ○ | ○ |
| Tightness or pressure in chest | ○ | ○ | ○ |
| Loss of or change in smell or taste | ○ | ○ | ○ |
| Lack of appetite like you just haven't been hungry | ○ | ○ | ○ |
| Abdominal pain | ○ | ○ | ○ |
| Nausea or vomiting | ○ | ○ | ○ |
| Diarrhea | ○ | ○ | ○ |
| Constipation | ○ | ○ | ○ |
| Reflux/GERD/heartburn | ○ | ○ | ○ |
| Joint pain | ○ | ○ | ○ |
| Muscle pain | ○ | ○ | ○ |
| Headaches | ○ | ○ | ○ |
| Problems thinking or concentrating ("brain fog") | ○ | ○ | ○ |
| Nerve problems (tremor, shaking, abnormal movements, numbness, tingling, burning, can't move part of body, new seizures) | ○ | ○ | ○ |
| Problems with anxiety, depression, stress, or trauma-related symptoms like nightmares or grief | ○ | ○ | ○ |
| Problems with sleep | ○ | ○ | ○ |

### Consent form

Please read the consent form. If you have any questions, you can reach us at or (206) 295-9238.

---

UNIVERSITY OF WASHINGTON

Consent to Participate in a Research Study Called:

homeRNA to evaluate early immune predictors of m-acute sequelae of COVID-19 (PASC)

Principal Investigator: Ashleigh B. Theberge, Ph.D., Assistant Professor, UW, Department of Chemistry

Study Coordinator: Filip Stefanovic, Ph.D. Student, UW, Department of Chemistry,

Jane Edelson, UW, Department of Chemistry,

This form gives you information to help you decide whether or not to be in the study. Being in the study is voluntary. Please read this carefully. You may ask any questions about the study. Then you can decide whether or not you want to participate.

---

What is the purpose of the study? You are being asked to volunteer for a pilot research study to help us understand more about the association between early immune response during infection and development of long COVID (also known as post-acute sequelae of COVID-19 (PASC) in underrepresented, understudied, and underreported women. In this study, you will collect blood and nasal samples from yourself. Blood samples will be collected with a user friendly, disposable, sterile, blood collection device (Tasso-SST); preserve the immune responses of blood cells, using the study's homeRNA kit; and mail samples back to the research lab. The Tasso-SST device used in this study is an investigational medical device, not approved by the United States Food and Drug Administration (FDA), that enables blood collection in the home setting.

What procedures are involved in the study? If you choose to participate in this study, study procedures may take place over up to 6 months. We will contact you by phone, text or email to schedule deliveries and will mail you kits for the self-collection of blood samples at home. During the study, we will discuss with you the timepoints to collect blood samples. You will be provided with an instructional pamphlet and video illustrating how to use the blood sampling kit, as well as an instructional pamphlet for the nasal swab procedure. The study team is available to answer questions by phone, text, email, or a video call to talk through these steps. Following completion of each kit, you will mail the sample collections to the study team laboratory, using special postage-paid envelopes we provide. Specific procedures of this study involve:

Initial enrollment survey: At the beginning of the study, we will send you a link to complete an initial survey online. The survey will ask you to provide information about health conditions you have, and your contact information, including mailing address. It will also ask questions about burdens you experience such as financial and obstacles accessing healthcare. It will take 15 minutes or less to complete this survey.

Self-collection blood draws: We will ask you to collect blood in 1 to 2 sets each consisting of 5 timepoints. If you complete the 1st set of samples, you will perform 7 blood draws at 5 timepoints over 5 weeks. You will perform 2 blood draws at timepoints 1 and 5 and 1 blood draw at timepoints 2,3,4. If you are asked to complete a second set of sample timepoints, you will perform 5 blood draws over an additional 8 weeks. We expect the timepoints for sampling will be specific to each person in the study. The study team will communicate the frequency with you over the study duration.

If you are asked to complete a second set of 5 sample timepoints, you will collect blood during additional timepoints determined by the study team. In this scenario, you would collect up to a total of 12 blood samples during the study. The maximum amount of blood you will collect over up to 13 weeks will not exceed 6 mL (about 1 teaspoon). However, if a blood sample gets lost in the mail or is otherwise unusable, we may ask for you to collect a replacement sample.

During sampling, you will collect blood from your upper arm using the Tasso-SST, following the instructions for use and information provided to you. You will push a button to deploy the device lancets, and the Tasso-SST device will remain on your arm for 2-3 minutes. The device will collect up to 10 drops of blood with each use (which is about 0.5 mL of blood, or less than 1/8 of a teaspoon). We expect it will take 15 minutes each time to draw the blood, stabilize it, and package it for return.

Self-Collected Nasal Swabs: At the same timepoints you collect blood, you will also collect 1-2 nasal samples. You will use a simple and comfortable method of collection to self-swab the interior of your ?nose. It will take about 5 minutes to perform a nasal swab at each time point. We will ask you to perforpost nasal swabs as follows:

1 set of sample timepoints: 5-10 nasal swabs over up to 5 weeks

If you are asked to complete a 2nd set of sample timepoints: 5-10 more nasal swabs over an additional 8 weeks.

The nasal swabs will be used to verify SARS-CoV-2 infection. Results will not be returned to you.

Surveys: For each day you perform blood and nasal sample collection, you will complete a brief online survey and provide information about you, your health information, your symptoms, stress levels, sleep, and other information. Some surveys will have questions about the device use and your general experience of the sample collection process during the study. You will also receive an additional online survey following the 5th and 10th blood and nasal sample collections that will ask about your experiences during the study. Each survey will take about 15 minutes or less to complete following sample collection. The additional surveys will take about 10 minutes to complete. You will also receive a survey at months 4, 5, and 6 following your enrollment in the study.

What are the risks of the study? Self-collection blood draw: The risks are similar to a finger stick blood collection test for measuring your blood sugar level. You may experience pain, bruising or bleeding at the site of the punctures during application of the Tasso-SST device. As with any blood draw, you may also feel faint or sick to your stomach while having your blood drawn. You should use the blood sampling device while sitting down to minimize the chance of fainting. There is a rare risk of infection due to the Tasso-SST. During the collection, you will stick the device to your upper arm. This may cause temporary discomfort where it is applied. The Tasso-SST collection device has been used in thousands of people and no severe complications have been reported.

Self-collection nasal swab: You may experience some mild discomfort, watery eyes, or sneezing.

Stress: It is possible that responding to surveys about symptoms related to your COVID-19 infection and PASC could cause you distress. Information we ask you to track is similar to how you might log your symptoms for your clinical care.

Privacy and confidentiality: There is a risk that the confidentiality of your participation in this study could be breached. We will do everything that we can to make sure that this does not happen.

There may be other risks of study participation that are unknown.

What happens if I have been injured because of this study?

It is important that you promptly inform the study staff if you believe you have been injured from the procedures you perform in this study. You can call the study team at the number(s) listed at the end of the form. The UW does not normally provide compensation for harm except through its discretionary program for medical injury. However, the law may allow you to seek other compensation if the harm is the fault of the researchers. You do not waive any right to seek payment by signing this consent form.

What are the benefits of this study? There may be no direct benefits to you. Your participation in this study will help us understand immune response in individuals with PASC (long COVID). Long term, we hope information from our study may aid in advancing precision treatment of PASC (long COVID).

---

Will I receive payment for this study? You will be paid in digital gift cards. You will be paid after completion of each month or following a set
of 5 samples (whichever comes first). If you withdraw from the study during a partial month, we will compensate you $10 per sample. Most participants will complete 5 timepoints and 3 follow-up surveys and will be compensated up to $160. A subset of participants will be invited for extended sampling and will collect and complete an additional 5 sample timepoints consisting of blood samples, nasal swabs, and sampling surveys as well as additional surveys about their study experience and will be compensated up to another $160.
Please find the compensation summary outlined in the table below:
Set 1: (Up to $160 for completion of Set 1)
  Timepoints 1-5: $20 each timepoint
  3 monthly symptom surveys after Timepoint 5: $20 per survey

Set 2: (Up to $130 for completion of Set 2)
  Timepoints 6-10: $20 each timepoint
  3 monthly symptom surveys after Timepoint 10: $20 per survey

How will the information about me be kept private? Your record will be kept confidential to the extent described here. Data and samples provided in this
study will be coded with a unique study number and the link between identifiers and the unique study number will be stored separately in a study database. You will not be identified in any report about this study. Researchers at the University of Washington will be able to see your data from the study in order to study the results and process your samples. Government and university staff sometimes review studies such as this one to make sure they are being done safely and legally. If a review of this study takes place, your study records may be examined. The reviewers will protect your privacy.

We have a Certificate of Confidentiality from the federal National Institutes of Health. This helps us protect your privacy. The Certificate means that we do not have to give out information, documents, or samples that could identify you even if we are asked to by a court of law. We will use the Certificate to resist any demands for identifying information.

We can't use the Certificate to withhold your research information if you give your written consent to give it to an insurer, employer, or other person. Also, you or a member of your family can share information about yourself or your part in this research if you wish.

There are some limits to this protection. We will voluntarily provide the information to:
- a member of the federal government who needs it in order to audit or evaluate the research;
- individuals at the institution(s) conducting the research, the funding agency, and other groups involved in the research, if they need the information to make sure the research is being done correctly;
- the federal Food and Drug Administration (FDA), if required by the FDA;
- individuals who want to conduct secondary research if allowed by federal regulations and according to your consent for future research use as described in this form;

The Certificate expires when the NIH funding for this study ends. Currently this is 7/31/2023. Any data collected after expiration is not protected as described above. Data collected prior to expiration will continue to be protected.

---

How is this study funded? The study team is receiving financial support from the National Institutes of Health, the David and
Lucile Packard Foundation, the Alfred P. Sloan Foundation, and by the University of Washington.

---

How will my information and samples be used for research? Using Your Samples and Data in Future Research: The information and/or specimens that we obtain from you for this study might be used for future studies. We may remove anything that might identify you from the information and specimens. If we do so, that information and specimens may then be used for future research studies or given to another investigator without getting additional permission from you. It is also possible that data used or shared for future research may be in an identifiable format. If we do, a review board will decide whether or not we need to get additional permission from you.

Commercial profit: The specimen we collect as part of this research may be used for commercial profit. There is no plan to share this profit with you.

---

Can I withdraw from this study? YYou may choose not to participate in the study at all. Your decision to participate is voluntary. You may
choose to withdraw from the study at any time, for any reason.

Please inform the study team right away if you become pregnant during your participation in the study. You will not be eligible to participate in the study for the duration of your pregnancy. You may express your wish to stop participating in the study by contacting the study staff at or (206) 295-9238. If you request to withdraw from the study, we will keep the data and samples collected to that point. You may be withdrawn if you miss two collection time points and do not respond to efforts by the study team.

Who can I call if I have questions or problems?

For Questions About Please Contact
This study (including complaints and requests for information) Filip Stefanovic at:

, (206)295-9238

Consent to Participant in this Research Study?

I have carefully read the description of this study. This study has been explained to me. I have a choice whether to take part in this study. I understand what is involved in being a volunteer. I have been told of the risks and benefits of being in this study. I have had the chance to ask questions about it, and all questions were answered to my satisfaction. If I have questions later about the research, or if I have been harmed by participating in this study, I can contact the study number listed above. If I have questions about my rights as a research subject, I can call the Human Subjects Division at (206) 543-0098 or call collect at (206) 221-5940. I do not give up any of my legal rights by signing this consent form. I now agree to take part in this research study.

A copy of the consent form will be emailed to you at an email address that you provide. It will be a "PDF" document. Most computers already have PDF viewer software installed, which will allow you to open, read, or print the consent form. The email we send you will include a link to PDF viewer software (such as Adobe Acrobat Reader) in case your computer does not already have it. If you would prefer to receive a paper copy of the consent form at no cost to you, please contact the researcher listed on page 4 of this consent form.

Research Participant's Printed Name:
(18+ years or older)

______ ______

Date: ______

Please sign below

1) Signature:

________________________________________

FUTURE RESEARCH CONTACT

If you are interested in returning for other studies that the Bioanalytical Chemistry for Medicine and the Environment (BCME) group operates, the BCME group could keep basic contact information in a private database. This would not be linked to the data collected in the different studies and you can be removed from this list at any time by request. The contact information will be securely stored and will not be sold. Please indicate below if you want to be re-contacted for further study recruitment:

______

### Welcome Survey

This survey will ask you question about your health.

#### About You and Your Health

What is your date of birth? MM/DD/YY

______________________________

What is your height?

Feet: ______ Inches: ______

What is your weight in pounds?

______________________________

Did you have any pre-existing medical conditions or experience any of the following before your current COVID-19 infection? (Please select all that apply)

- ☐ Asthma
- ☐ Seasonal allergies
- ☐ Congestive heart failure
- ☐ High blood pressure
- ☐ High cholesterol
- ☐ Irritable bowel syndrome (IBS)
- ☐ Chronic constipation
- ☐ Diarrhea
- ☐ Liver disease
- ☐ Colitis
- ☐ Reflux
- ☐ Systemic lupus erythematosus (SLE)
- ☐ Multiple sclerosis (MS)
- ☐ Rheumatoid arthritis (RA)
- ☐ Thyroid disease
- ☐ Diabetes (type I, type II, or pre-diabetic)
- ☐ Depression
- ☐ Anxiety
- ☐ Cancer
- ☐ Kidney disease
- ☐ None of the above
- ☐ Other (please specify)

Please indicate other pre-existing conditions/diagnoses not listed here. If multiple, please separate them with a comma. Please only list the conditions, no descriptions or explanations.

______________________________

#### Have you previously been tested for any of these conditions? If so, please indicate when and the test results:

| | Not tested | Negative | Current/recent infection (last year) | Past infection (more than a year ago) | Not sure |
|---|---|---|---|---|---|
| Epstein-Barr (mono) | ○ | ○ | ○ | ○ | ○ |
| Lyme disease | ○ | ○ | ○ | ○ | ○ |
| Cytomegalovirus (CMV) | ○ | ○ | ○ | ○ | ○ |

| | |
|---|---|
| Have any of your pre-existing conditions changed since your COVID-19 diagnosis? | ◯ Yes, they got worse<br>◯ Yes, they got better<br>◯ Some got better, some stayed the same, some got worse (please add an explanation in the text boxes in the following page)<br>◯ No, they stayed the same<br>◯ N/A (I did not have a pre-existing condition) |
| Did you receive a new medical diagnosis related to COVID-19? | ◯ Yes<br>◯ No |
| Have you been newly diagnosed with any illness or complication related to COVID-19? (select all that apply) | ☐ Cardiovascular<br>☐ Dermatological<br>☐ Endocrine<br>☐ Gastro-intestinal<br>☐ Musculoskeletal<br>☐ Mental Health<br>☐ Neurological<br>☐ Pulmonary<br>☐ Renal<br>☐ Other |
| Please describe any other diagnosis you were given (if multiple, please put each diagnosis on a new line by clicking enter/return) | ______________________________ |
| Did you require hospitalization as a result of your current COVID-19 infection? | ◯ Yes<br>◯ No |
| If you were hospitalized, approximately for how long? | ________________________<br>(In days) |
| Do you currently take any medications regularly? | ◯ Yes<br>◯ No |
| If yes, please list out the medications and frequency of use here:<br>(enter each medication and its frequency on a new line by pressing enter/return) | ______________________________ |
| Have you taken Paxlovid to treat your current COVID-19 infection? | ◯ Yes<br>◯ No |
| Have you taken any other medication or supplement to treat your current COVID-19 infection? | ◯ Yes<br>◯ No |
| Please describe the type and frequency of substance used:<br>(put each new medication or supplement on a new line by clicking enter/return) | ______________________________ |
| Do you have a permanent or temporary disability? | ◯ Yes<br>◯ No |
| If so, please briefly describe your disability/disabilities here: | ______________________________ |

What is your blood type? If you don't know, please select 'Don't know'.

◯ A+
◯ A-
◯ B+
◯ B-
◯ AB+
◯ AB-
◯ O+
◯ O-
◯ Don't Know

Please select all listed symptom(s) below that you are experiencing today.

Nose Symptoms
Runny nose ______
Congestion ______
Sinus pain ______
Nasal drip ______
Sneezing ______
Ears Symptoms
Ear pain ______
Throat Symptoms
Sore throat ______
Hoarseness ______
Chest Symptoms
Watery or itchy eyes ______
Cough ______
Phlegm or mucous production ______
Wheezing or chest tightness ______
Shortness of breath ______
Chest pain ______
Racing heart/palpitations/arrhythmia ______
Gastrointestinal Symptoms
Diarrhea ______
Nausea ______
Abdominal pain ______
Vomiting ______
Constipation ______
Reflux/GERD/heartburn ______
General Symptoms
Fatigue ______
Feeling fatigued after exercise ______
Dizziness ______
Fever ______
Chills ______
Headache ______
Aching muscles ______
Joint pain ______
Sleep Changes Symptoms
Sleep disruption/insomnia ______
Sensory Changes Symptoms
Change in or loss of smell ______
Change in or loss of taste ______
Mental Health Symptoms
Anxiety ______
Depression ______
Stress ______

#### Physical Health, Mental Health, Lifestyle

Do you currently or have you ever uses any tobacco products?

◯ Yes
◯ No

Which of the following tobacco products do you or have you used?

☐ cigarettes
☐ e-cigarettes
☐ vape
☐ pipes
☐ cigars
☐ smokeless cigarettes

How many cigarettes do you or used to smoke per day on average?

__________________________________
(Note: there are 20 cigarettes in a pack)

How many years have you smoked cigarettes?

__________________________________

Have you quit smoking for a period of time?

◯ Yes
◯ No

Please describe how long you quit cigarettes for?

__________________________________________

Please specify the use frequency and quantity of other tobacco products (not cigarettes):

__________________________________________

Did you receive a flu vaccination in the last year?

◯ Yes
◯ No

Do you currently take any non-steroidal anti-inflammatory agents (NSAIDS) at least once a week, with or without a prescription?

◯ Yes
◯ No
(These include: ibuprofen (Motrin, Advil) naproxen (Naprosyn, Alene, Anaprox, Naprelan) diclofenac (Cambia, Catalan, Voltaire's, Zipsor) indomethacin (Indocin) diflunisal (Dolobid) etodolac (Lodine, Lodine XL) ketorolac (Acular, Acular LS, Acular PF, Acuvail) nambumetone (Relafen) oxaprozin (Daypro) piroxicam (Feldene) salsalate (Disalate) sulindac (Clinoril) tormenting (Tolectin 600, Tolectin DS) celecoxib (Cerebrex)))

Do you take aspirin at least once a week, with or without a prescription?

◯ Yes
◯ No

Do you currently take any other medications (including over the counter medications) at least once a week?

◯ Yes
◯ No

Please list these medications/supplements and the frequency of use here (separate each entry with a comma)

__________________________________________
(e.g., turmeric supplements (once per day), vitamin c (once a week))

Overall, how satisfied are you with life as a whole these days?
(0 = Not Satisfied at All, 10 = Completely Satisfied )

◯ 0 ◯ 1 ◯ 2 ◯ 3 ◯ 4 ◯ 5 ◯ 6 ◯ 7 ◯ 8 ◯ 9 ◯ 10

In general, how happy or unhappy do you usually feel?
(0 = Extremely Unhappy, 10 = Extremely Happy)

◯ 0 ◯ 1 ◯ 2 ◯ 3 ◯ 4 ◯ 5 ◯ 6 ◯ 7 ◯ 8 ◯ 9 ◯ 10

In general, how would you rate your physical health?
(0 = Poor, 10 = Excellent)

◯ 0 ◯ 1 ◯ 2 ◯ 3 ◯ 4 ◯ 5 ◯ 6 ◯ 7 ◯ 8 ◯ 9 ◯ 10

How would you rate your overall mental health?
(0 = Poor, 10 = Excellent )

◯ 0 ◯ 1 ◯ 2 ◯ 3 ◯ 4 ◯ 5 ◯ 6 ◯ 7 ◯ 8 ◯ 9 ◯ 10

I am content with my friendships and relationships.
(0 = Strongly Disagree, 10 = Strongly Agree )

◯ 0 ◯ 1 ◯ 2 ◯ 3 ◯ 4 ◯ 5 ◯ 6 ◯ 7 ◯ 8 ◯ 9 ◯ 10

My relationships are as satisfying as I would want them to be.
(0 = Strongly Disagree, 10 = Strongly Agree )

◯ 0 ◯ 1 ◯ 2 ◯ 3 ◯ 4 ◯ 5 ◯ 6 ◯ 7 ◯ 8 ◯ 9 ◯ 10

How often do you worry about being able to meet normal monthly living expenses?
(0 = Worry All of the Time, 10 = Do Not Ever Worry)

◯ 0 ◯ 1 ◯ 2 ◯ 3 ◯ 4 ◯ 5 ◯ 6 ◯ 7 ◯ 8 ◯ 9 ◯ 10

How often do you worry about safety, food, or housing?
(0 = Worry All of the Time, 10 = Do Not Ever Worry)

◯ 0 ◯ 1 ◯ 2 ◯ 3 ◯ 4 ◯ 5 ◯ 6 ◯ 7 ◯ 8 ◯ 9 ◯ 10

#### Background, Demographics, Access to Healthcare

Do you have children under the age of 18 at home?

◯ Yes
◯ No

Are you the primary caregiver for a parent or elderly person in your household?

◯ Yes
◯ No

What is the highest grade or level of school you have completed or the highest degree you have received?

◯ Less than high school
◯ High school diploma or equivalent (GED)
◯ Associate degree
◯ Bachelor's degree or 4-year college
◯ Master's degree
◯ Doctoral degree

What is your occupation?

________________________________

| | |
|---|---|
| What is your marital status? | ◯ Single<br>◯ Long-term partner<br>◯ Married<br>◯ Separated<br>◯ Divorced<br>◯ Widowed |
| Are you currently covered by health insurance? | ◯ Yes<br>◯ No |
| When was the last time you had a wellness checkup or physical? | ◯ < 1 year ago<br>◯ 1-2 years ago<br>◯ 2-3 years ago<br>◯ >3 years ago |
| If you need healthcare, how far do you have to travel to be seen? Note that this isn't necessarily your closest hospital/clinic. | ◯ < 1 mile<br>◯ 1-2 miles<br>◯ 2-3 miles<br>◯ 4-5 miles<br>◯ 5-7 miles<br>◯ 7-10 miles<br>◯ >10 miles |
| Do you go to any of these places when you are sick and need health care? | ◯ Doctor's office or health center<br>◯ Urgent care center<br>◯ Clinic in a drug store or grocery store<br>◯ Hospital emergency room<br>◯ VA Medical Center<br>◯ VA outpatient clinic<br>◯ Telehealth (Zoom or other online format)<br>◯ Some other place<br>◯ I do not seek health care even if it is needed |
| During the past 12 months, what is the total combined number of times you visited those places with a health related matter? ex. 3 (integer only) | ________________________________ |
| How often do you not seek medical attention when you need it? | ◯ Never<br>◯ Sometimes<br>◯ Most of the time<br>◯ All of the time |
| What prevents you from seeking medical attention? (select all that apply) | ☐ Lack of or inadequate health insurance<br>☐ Not enough time/too busy<br>☐ Inconvenient clinic hours (i.e. during work hours)<br>☐ Transportation too difficult<br>☐ Lack of childcare<br>☐ Language barriers<br>☐ Do not trust doctors<br>☐ Other |
| What is your estimated average household yearly income? | ◯ < $50,000<br>◯ $50-75,000<br>◯ $75-100,000<br>◯ >$100,000<br>◯ Prefer not to answer |

| | |
|---|---|
| How many months of living expenses can you cover with your current savings? | ◯ No months<br>◯ Less than 1 month<br>◯ 1 to 3 months<br>◯ 4 to 6 months<br>◯ More than 6 months<br>◯ Prefer not to say |
| The amount of debt I have overwhelms me | ◯ Yes<br>◯ No |

### Mailing Address for Delivery of Study Kit

Please provide the mailing address at which you would like to receive your study kit.

---

Study Kit Delivery: What address would you like to receive your kit?
(Please do not include P.O. Boxes)

Street address:
(include unit or apartment number)

______
City: ______
State: ______
Zip Code: ______

Please indicate any special delivery instructions for receiving your study kit.

______

---

Study Kit Return: Is the pickup address different than the delivery address?

○ Yes ○ No

---

Pick up address (if different from delivery address)
(Please do not include P.O. Boxes)

Street address:
(include unit or apartment number)

______
City: ______
State: ______
Zip Code: ______

Please provide specific instructions for pickup of your completed test kits.

______

---

Study Kit Return: Please choose where you will leave your completed kit to be picked up.

○ Front Door/Porch
○ Back Door/Porch
○ Garage
○ Reception/Lobby
○ Office
○ Mailroom

### Sample Survey Sample Point #1

#### Sampling Survey

Please select all listed symptom(s) below that you are experiencing today.

Nose Symptoms
Runny nose ______
Congestion ______
Sinus pain ______
Nasal drip ______
Sneezing ______
Ears Symptoms
Ear pain ______
Throat Symptoms
Sore throat ______
Hoarseness ______
Chest Symptoms
Watery or itchy eyes ______
Cough ______
Phlegm or mucous production ______
Wheezing or chest tightness ______
Shortness of breath ______
Chest pain ______
Racing heart/palpitations/arrhythmia ______
Gastrointestinal Symptoms
Diarrhea ______
Nausea ______
Abdominal pain ______
Vomiting ______
Constipation ______
Reflux/GERD/heartburn ______
General Symptoms
Fatigue ______
Feeling fatigued after exercise ______
Dizziness ______
Fever ______
Chills ______
Headache ______
Aching muscles ______
Joint pain ______
Sleep Changes Symptoms
Sleep disruption/insomnia ______
Sensory Changes Symptoms
Change in or loss of smell ______
Change in or loss of taste ______
Mental Health Symptoms
Anxiety ______
Depression ______
Stress ______

#### Device Use Survey

homeRNA blood kit code:

__________________________________
(e.g., UU-XX-B-1)

homeRNA blood kit time:

__________________________________

What is the temperature and humidity reading on your thermometer today?

Temperature (?F) ______
Humidity (%) ______

#### About your experience when using the homeRNA blood kit today:

Aproximately how long (in minutes) did it take you to use the homeRNA blood kit today?

- ◯ Less than 5 minutes
- ◯ 5-10 minutes
- ◯ 11-15 minutes
- ◯ 16-20 minutes
- ◯ More than 20 minutes

While using the homeRNA blood kit today, approximately how long (in minutes) did you leave the Tasso-SST blood collection device on your arm?

- ◯ Less than 3 minutes
- ◯ 3-5 minutes
- ◯ More than 5 minutes
- ◯ I am unsure

Based on the image below, how much blood did you roughly collect today? Choose the closest Level.

- ◯ I was unable to collect any blood
- ◯ Level 1
- ◯ Level 2
- ◯ Level 3
- ◯ Level 4

______

| | No pain | Mild pain | Moderate pain | Severe pain | Very severe pain |
|---|---|---|---|---|---|
| Did you experience any pain when using the homeRNA blood kit today? | ◯ | ◯ | ◯ | ◯ | ◯ |

Has the temporary pain resolved?

- ◯ Yes
- ◯ No

We want to take a moment to sincerely apologize for any pain or discomfort that you may have experienced during the study procedures. Our team is committed to ensuring the well-being of our participants, and we take your feedback seriously. We encourage you to reach out to us if you need any assistance or would like to discuss your experience further. Phone number: (206)295-9238

Did you experience any issues with the Tasso-SST blood collection device?

- ◯ Yes
- ◯ No

Please describe any issues you had with the Tasso-SST blood collection device.

Did you experience any issues with the stabilizer tube or mixing your samples?

- ◯ Yes
- ◯ No

Please describe any issues you had with the stabilizer tube or mixing your samples. (Optional)

Please use this space for any other comments on your experience today collecting blood samples using the homeRNA blood kit (optional).

#### General Check-in

| | |
|---|---|
| Have your medications changed since you last filled out a survey? | ☐ I started taking a new medication<br>☐ I stopped taking a medication<br>☐ My medications have not changed since I last filled out a survey |
| Please describe the new medication: | __________________________________ |
| Please describe which medication you stopped taking: | __________________________________ |
| Has your address changed since you last filled out a survey? | ◯ Yes<br>◯ No |

### Sample Survey Sample Point #2

#### Sampling Survey

Please select all listed symptom(s) below that you are experiencing today.

Nose Symptoms
Runny nose ______
Congestion ______
Sinus pain ______
Nasal drip ______
Sneezing ______
Ears Symptoms
Ear pain ______
Throat Symptoms
Sore throat ______
Hoarseness ______
Chest Symptoms
Watery or itchy eyes ______
Cough ______
Phlegm or mucous production ______
Wheezing or chest tightness ______
Shortness of breath ______
Chest pain ______
Racing heart/palpitations/arrhythmia ______
Gastrointestinal Symptoms
Diarrhea ______
Nausea ______
Abdominal pain ______
Vomiting ______
Constipation ______
Reflux/GERD/heartburn ______
General Symptoms
Fatigue ______
Feeling fatigued after exercise ______
Dizziness ______
Fever ______
Chills ______
Headache ______
Aching muscles ______
Joint pain ______
Sleep Changes Symptoms
Sleep disruption/insomnia ______
Sensory Changes Symptoms
Change in or loss of smell ______
Change in or loss of taste ______
Mental Health Symptoms
Anxiety ______
Depression ______
Stress ______

#### Device Use Survey

homeRNA blood kit code:

__________________________________
(e.g., UU-XX-B-1)

homeRNA blood kit time:

__________________________________

What is the temperature and humidity reading on your thermometer today?

Temperature (?F) ______
Humidity (%) ______

#### About your experience when using the homeRNA blood kit today:

Aproximately how long (in minutes) did it take you to use the homeRNA blood kit today?

- ◯ Less than 5 minutes
- ◯ 5-10 minutes
- ◯ 11-15 minutes
- ◯ 16-20 minutes
- ◯ More than 20 minutes

While using the homeRNA blood kit today, approximately how long (in minutes) did you leave the Tasso-SST blood collection device on your arm?

- ◯ Less than 3 minutes
- ◯ 3-5 minutes
- ◯ More than 5 minutes
- ◯ I am unsure

Based on the image below, how much blood did you roughly collect today? Choose the closest Level.

- ◯ I was unable to collect any blood
- ◯ Level 1
- ◯ Level 2
- ◯ Level 3
- ◯ Level 4

______

| | No pain | Mild pain | Moderate pain | Severe pain | Very severe pain |
|---|---|---|---|---|---|
| Did you experience any pain when using the homeRNA blood kit today? | ◯ | ◯ | ◯ | ◯ | ◯ |

Has the temporary pain resolved?

- ◯ Yes
- ◯ No

We want to take a moment to sincerely apologize for any pain or discomfort that you may have experienced during the study procedures. Our team is committed to ensuring the well-being of our participants, and we take your feedback seriously. We encourage you to reach out to us if you need any assistance or would like to discuss your experience further. Phone number: (206)295-9238

Did you experience any issues with the Tasso-SST blood collection device?

- ◯ Yes
- ◯ No

Please describe any issues you had with the Tasso-SST blood collection device.

Did you experience any issues with the stabilizer tube or mixing your samples?

- ◯ Yes
- ◯ No

Please describe any issues you had with the stabilizer tube or mixing your samples. (Optional)

Please use this space for any other comments on your experience today collecting blood samples using the homeRNA blood kit (optional).

#### General Check-in

| | |
|---|---|
| Have your medications changed since you last filled out a survey? | ☐ I started taking a new medication<br>☐ I stopped taking a medication<br>☐ My medications have not changed since I last filled out a survey |
| Please describe the new medication: | __________________________________ |
| Please describe which medication you stopped taking: | __________________________________ |
| Has your address changed since you last filled out a survey? | ◯ Yes<br>◯ No |

### Closing Survey

#### Closing Survey

General Check-In

Overall, how satisfied are you with life as a whole these days?
(0 = Not Satisfied at All, 10 = Completely Satisfied )

○ 0 ○ 1 ○ 2 ○ 3 ○ 4 ○ 5 ○ 6 ○ 7 ○ 8 ○ 9 ○ 10

In general, how happy or unhappy do you usually feel?
(0 = Extremely Unhappy, 10 = Extremely Happy)

○ 0 ○ 1 ○ 2 ○ 3 ○ 4 ○ 5 ○ 6 ○ 7 ○ 8 ○ 9 ○ 10

In general, how would you rate your physical health?
(0 = Poor, 10 = Excellent)

○ 0 ○ 1 ○ 2 ○ 3 ○ 4 ○ 5 ○ 6 ○ 7 ○ 8 ○ 9 ○ 10

How would you rate your overall mental health?
(0 = Poor, 10 = Excellent )

○ 0 ○ 1 ○ 2 ○ 3 ○ 4 ○ 5 ○ 6 ○ 7 ○ 8 ○ 9 ○ 10

I am content with my friendships and relationships.
(0 = Strongly Disagree, 10 = Strongly Agree )

○ 0 ○ 1 ○ 2 ○ 3 ○ 4 ○ 5 ○ 6 ○ 7 ○ 8 ○ 9 ○ 10

My relationships are as satisfying as I would want them to be.
(0 = Strongly Disagree, 10 = Strongly Agree )

○ 0 ○ 1 ○ 2 ○ 3 ○ 4 ○ 5 ○ 6 ○ 7 ○ 8 ○ 9 ○ 10

How often do you worry about being able to meet normal monthly living expenses?
(0 = Worry All of the Time, 10 = Do Not Ever Worry)

○ 0 ○ 1 ○ 2 ○ 3 ○ 4 ○ 5 ○ 6 ○ 7 ○ 8 ○ 9 ○ 10

How often do you worry about safety, food, or housing?
(0 = Worry All of the Time, 10 = Do Not Ever Worry)

○ 0 ○ 1 ○ 2 ○ 3 ○ 4 ○ 5 ○ 6 ○ 7 ○ 8 ○ 9 ○ 10

Overall Study Experience

Please rate your overall experience in the study on a scale from 1 to 5 with 1 being poor and 5 being excellent

○ 1 ○ 2 ○ 3 ○ 4 ○ 5

On a scale of 1 to 5 (5 being very easy and 1 being very challenging), how easy was it for you to do the blood/nasal sampling and surveys with your daily routine and other responsibilities (like work or caring for others)?

◯ 1 ◯ 2 ◯ 3 ◯ 4 ◯ 5

| | |
|---|---|
| Is there anything we can do to make it easier for you in the future? | ◯ Yes<br>◯ No |
| Please describe how we can make this process easier for future participants: | __________________________________ |
| Have you participated in research studies previously? | ◯ Yes<br>◯ No |
| Were they remote/at-home like this study or in person studies (requiring visiting a hospital, clinic, or research site)? | ☐ Remote/at-home study<br>☐ In person study<br>(can select both if applicable) |
| Please describe the nature of the study (survey only, survey + blood/urine/other sample) | ___________________________ |
| Based your experience in this study, would you participate in a similar remote or at-home study in the future? | ◯ Yes<br>◯ No |
| Do you think there will be any challenges for you to participate in similar studies in the future? | ◯ Yes<br>◯ No |
| If so, please briefly describe these challenges: | __________________________________ |
| Would you participate in an in-person study requiring visiting a hospital, clinic, or research site in the future? | ◯ Yes<br>◯ No |
| If so, do you foresee any challenges to participating in an in-person study? | ◯ Yes<br>◯ No |
| If so, please briefly describe these challenges: | __________________________________ |
| Receiving the Kit | |
| Did you receive your kit by the expected time? | ◯ Yes<br>◯ No |
| If No, Please Explain (optional). | |
| Was there any visible damage to the box or components? | ◯ Yes<br>◯ No |

| | |
|---|---|
| If Yes, can you describe the damage(s)? (optional) | __________________________________ |
| Did you experience any issues in receiving your kit? | ◯ Yes<br>◯ No |
| If Yes, can you describe the issue(s)? (optional) | __________________________________ |

#### Using the Kit

**How easy was it to use the homeRNA kit overall?**

| | Very easy | Somewhat easy | Neither easy nor hard | Somewhat hard | Very hard |
|---|---|---|---|---|---|
| Tasso-SST device | ○ | ○ | ○ | ○ | ○ |
| Stabilization tube | ○ | ○ | ○ | ○ | ○ |

**How easy was it to use the Nasal swab kit?**

| | Very easy | Somewhat easy | Neither easy nor hard | Somewhat hard | Very hard |
|---|---|---|---|---|---|
| Swab collection | ◯ | ◯ | ◯ | ◯ | ◯ |
| Transport tube | ◯ | ◯ | ◯ | ◯ | ◯ |

Can you please describe any part of the Instructions for use that was confusing or challenging to understand? (optional)

Were you able to use the kits and complete the collections within the designated time window?
◯ Yes
◯ No

Returning the Kits

If No, please describe what can be improved to allow you to use the kits within the designated time window.

Did you have problem(s) packaging the kit components?
◯ Yes
◯ No

If Yes, please describe the problem(s) you had.

Were the instructions for packaging and returning the kits clear?
◯ Yes
◯ No

If No, please describe what can be improved.

Did you have problem(s) filling out or accessing this survey?
◯ Yes
◯ No

If Yes, please describe the problem(s) you had.

Please use this space for any other comments on using and returning the kits.

Closing Remarks

| | |
|---|---|
| Is there anything else you would like to share with us about your experience in the study? | ◯ Yes<br>◯ No |
| Please use the space provided to share your feedback here: | __________________________________ |

### U3 Closing Survey

#### Closing Survey

General Check-In

Overall, how satisfied are you with life as a whole these days?
(0 = Not Satisfied at All, 10 = Completely Satisfied )

◯ 0 ◯ 1 ◯ 2 ◯ 3 ◯ 4 ◯ 5 ◯ 6 ◯ 7 ◯ 8 ◯ 9 ◯ 10

In general, how happy or unhappy do you usually feel?
(0 = Not Satisfied at All, 10 = Completely Satisfied )

◯ 0 ◯ 1 ◯ 2 ◯ 3 ◯ 4 ◯ 5 ◯ 6 ◯ 7 ◯ 8 ◯ 9 ◯ 10

In general, how would you rate your physical health?
(0 = Poor, 10 = Excellent)

◯ 0 ◯ 1 ◯ 2 ◯ 3 ◯ 4 ◯ 5 ◯ 6 ◯ 7 ◯ 8 ◯ 9 ◯ 10

How would you rate your overall mental health?
(0 = Poor, 10 = Excellent)

◯ 0 ◯ 1 ◯ 2 ◯ 3 ◯ 4 ◯ 5 ◯ 6 ◯ 7 ◯ 8 ◯ 9 ◯ 10

I am content with my friendships and relationships.
(0 = Strongly Disagree, 10 = Strongly Agree )

◯ 0 ◯ 1 ◯ 2 ◯ 3 ◯ 4 ◯ 5 ◯ 6 ◯ 7 ◯ 8 ◯ 9 ◯ 10

My relationships are as satisfying as I would want them to be.
(0 = Strongly Disagree, 10 = Strongly Agree )

◯ 0 ◯ 1 ◯ 2 ◯ 3 ◯ 4 ◯ 5 ◯ 6 ◯ 7 ◯ 8 ◯ 9 ◯ 10

How often do you worry about being able to meet normal monthly living expenses?
(0 = Worry All of the Time, 10 = Do Not Ever Worry)

◯ 0 ◯ 1 ◯ 2 ◯ 3 ◯ 4 ◯ 5 ◯ 6 ◯ 7 ◯ 8 ◯ 9 ◯ 10

How often do you worry about safety, food, or housing?
(0 = Worry All of the Time, 10 = Do Not Ever Worry)

◯ 0 ◯ 1 ◯ 2 ◯ 3 ◯ 4 ◯ 5 ◯ 6 ◯ 7 ◯ 8 ◯ 9 ◯ 10

What is your estimated average household yearly income?

◯ < $50,000
◯ $50-75,000
◯ $75-100,000
◯ >$100,000
◯ Prefer not to answer

Overall Study Experience

We sent two extra blood collection devices. How often did you use your extra Tasso-STT blood collection device?

- ◯ Always
- ◯ Often
- ◯ Sometimes
- ◯ Rarely
- ◯ Never

Which of the following motivated you to enroll in the study (please check all that apply)?

- ☐ An interest in adding to the body of knowledge on the health impacts of COVID-19
- ☐ The opportunity to expand representation of people like myself in scientific research
- ☐ A general interest in science
- ☐ The financial compensation I received for participating in the study
- ☐ The flexibility of a remote study
- ☐ Other motivation not listed above (Please explain:________)

Please specify

__________________________________

Which of the following motivated you to continue to participate in the study after you had enrolled (please check all that apply)?

- ☐ An interest in adding to the body of knowledge on the health impacts of COVID-19
- ☐ The opportunity to expand representation of people like myself in scientific research
- ☐ A general interest in science
- ☐ The financial compensation I received for participating in the study
- ☐ Positive interactions with the study team
- ☐ The flexibility of a remote study
- ☐ Other motivation not listed above (Please explain:________)

Please specify

__________________________________

Compared to participating in a study that requires in person visits, how easy would it be to participate in a remote study with multiple samples?

- ◯ Remote blood sampling would be significantly easier
- ◯ Remote blood sampling would be somewhat easier
- ◯ Remote blood sampling would be neither easier nor harder than in-person
- ◯ Remote blood sampling would be somewhat harder
- ◯ Remote blood sampling would be significantly harder

Please briefly describe why you selected your answer above:

__________________________________________

Have you participated in an in-person blood sampling study (e.g., having your blood drawn at a blood draw facility or clinic for the purpose of a research study) previously?

- ◯ Yes
- ◯ No
- ◯ Unsure

| | |
|---|---|
| Would any of the following prevent you from participating in an in-person blood sampling study (please check all that apply)? | ☐ Length of commute to the nearest clinic or blood draw facility<br>☐ Difficulty obtaining transportation to the clinic or blood draw facility<br>☐ Difficulty fitting blood draw visits into a work schedule or caregiver schedule<br>☐ Discomfort with the method of drawing blood at clinics or blood draw facilities (e.g., using a syringe or collection needle to draw blood from the inner elbow)<br>☐ Discomfort with going to clinics or other medical facilities<br>☐ Other (Please explain:_________)<br>☐ None of the above |

**Please rate your level of agreement with the following statements (1= strongly disagree, 5= strongly agree).**

| | 1 | 2 | 3 | 4 | 5 |
|---|---|---|---|---|---|
| I enjoyed participating in this study | ○ | ○ | ○ | ○ | ○ |
| The instructions on how to use the Tasso-STT blood collection device within the kit were easy to follow. | ○ | ○ | ○ | ○ | ○ |
| The instructions on how to use the stabilizer tube (the tube that you screw together with the blood tube) within the kit were easy to follow. | ○ | ○ | ○ | ○ | ○ |
| The instructions and communications from the study team were easy to follow (e.g., where to drop off my blood samples for pickup) | ○ | ○ | ○ | ○ | ○ |
| The Tasso-SST was easy to use | ○ | ○ | ○ | ○ | ○ |
| The stabilizer tube was easy to use | ○ | ○ | ○ | ○ | ○ |
| It was easy to follow the sampling schedule | ○ | ○ | ○ | ○ | ○ |
| Accessing and filling out the surveys after each sampling was easy | ○ | ○ | ○ | ○ | ○ |
| A phone call after I enrolled in the study that explained the steps of study participation was helpful. | ○ | ○ | ○ | ○ | ○ |
| The flexibility in when I could take my blood sample (e.g., at the time of day that was most convenient) was an important factor in my ability to participate in this study. | ○ | ○ | ○ | ○ | ○ |

Please use the space below if you would like to give any additional feedback about the question above: ________________________________________

In the current study, communication from the study team was sent to you over email. If you were to participate in the study again, how would you prefer to receive information for the study team? (select all that apply)

- ☐ Email only
- ☐ Text message only
- ☐ Both email and text message
- ☐ Other (Please describe:_________)

Please specify: ___________________________________

| | |
|---|---|
| Did you watch the video on how to use the blood collection device and stabilization kit? | ◯ Yes<br>◯ No<br>◯ Unsure |
| The video was helpful. (1= strongly disagree, 5= strongly agree). | ◯ 1<br>◯ 2<br>◯ 3<br>◯ 4<br>◯ 5 |
| Would you be willing to participate in this study or a similar study again? | ◯ Yes<br>◯ No<br>◯ Unsure |
| Please indicate the maximum number of years that you would be willing to participate in a similar remote blood sampling study assuming you were collecting ~15 samples per year: | ◯ 1 year<br>◯ 2 years<br>◯ 3 years<br>◯ 4 years |
| Is there anything we could change so that you would want to participate in this study or a similar remote blood sampling study again? | ______________________________ |
| Are there any other thoughts that you would like to share with the study team? | ______________________________ |

### Sample Survey Extended Sample Point #1

#### Sampling Survey

Please select all listed symptom(s) below that you are experiencing today.

Nose Symptoms
Runny nose ______
Congestion ______
Sinus pain ______
Nasal drip ______
Sneezing ______
Ears Symptoms
Ear pain ______
Throat Symptoms
Sore throat ______
Hoarseness ______
Chest Symptoms
Watery or itchy eyes ______
Cough ______
Phlegm or mucous production ______
Wheezing or chest tightness ______
Shortness of breath ______
Chest pain ______
Racing heart/palpitations/arrhythmia ______
Gastrointestinal Symptoms
Diarrhea ______
Nausea ______
Abdominal pain ______
Vomiting ______
Constipation ______
Reflux/GERD/heartburn ______
General Symptoms
Fatigue ______
Feeling fatigued after exercise ______
Dizziness ______
Fever ______
Chills ______
Headache ______
Aching muscles ______
Joint pain ______
Sleep Changes Symptoms
Sleep disruption/insomnia ______
Sensory Changes Symptoms
Change in or loss of smell ______
Change in or loss of taste ______
Mental Health Symptoms
Anxiety ______
Depression ______
Stress ______

#### Device Use Survey

homeRNA blood kit code:

__________________________________
(e.g., UU-XX-B-1)

homeRNA blood kit time:

__________________________________

What is the temperature and humidity reading on your thermometer today?

Temperature (?F) ______
Humidity (%) ______

#### About your experience when using the homeRNA blood kit today:

Aproximately how long (in minutes) did it take you to use the homeRNA blood kit today?

- ◯ Less than 5 minutes
- ◯ 5-10 minutes
- ◯ 11-15 minutes
- ◯ 16-20 minutes
- ◯ More than 20 minutes

While using the homeRNA blood kit today, approximately how long (in minutes) did you leave the Tasso-SST blood collection device on your arm?

- ◯ Less than 3 minutes
- ◯ 3-5 minutes
- ◯ More than 5 minutes
- ◯ I am unsure

Based on the image below, how much blood did you roughly collect today? Choose the closest Level.

- ◯ I was unable to collect any blood
- ◯ Level 1
- ◯ Level 2
- ◯ Level 3
- ◯ Level 4

______

| | No pain | Mild pain | Moderate pain | Severe pain | Very severe pain |
|---|---|---|---|---|---|
| Did you experience any pain when using the homeRNA blood kit today? | ◯ | ◯ | ◯ | ◯ | ◯ |

Has the temporary pain resolved?

- ◯ Yes
- ◯ No

We want to take a moment to sincerely apologize for any pain or discomfort that you may have experienced during the study procedures. Our team is committed to ensuring the well-being of our participants, and we take your feedback seriously. We encourage you to reach out to us if you need any assistance or would like to discuss your experience further. Phone number: (206)295-9238

Did you experience any issues with the Tasso-SST blood collection device?

- ◯ Yes
- ◯ No

Please describe any issues you had with the Tasso-SST blood collection device.

Did you experience any issues with the stabilizer tube or mixing your samples?

- ◯ Yes
- ◯ No

Please describe any issues you had with the stabilizer tube or mixing your samples. (Optional)

Please use this space for any other comments on your experience today collecting blood samples using the homeRNA blood kit (optional).

#### General Check-in

| | |
|---|---|
| Have your medications changed since you last filled out a survey? | ☐ I started taking a new medication<br>☐ I stopped taking a medication<br>☐ My medications have not changed since I last filled out a survey |
| Please describe the new medication: | __________________________________ |
| Please describe which medication you stopped taking: | __________________________________ |
| Has your address changed since you last filled out a survey? | ○ Yes<br>○ No |

### Extended Closing Survey

#### Closing Survey

General Check-In

Overall, how satisfied are you with life as a whole these days?
(0 = Not Satisfied at All, 10 = Completely Satisfied )

◯ 0 ◯ 1 ◯ 2 ◯ 3 ◯ 4 ◯ 5 ◯ 6 ◯ 7 ◯ 8 ◯ 9 ◯ 10

In general, how happy or unhappy do you usually feel?
(0 = Extremely Unhappy, 10 = Extremely Happy)

◯ 0 ◯ 1 ◯ 2 ◯ 3 ◯ 4 ◯ 5 ◯ 6 ◯ 7 ◯ 8 ◯ 9 ◯ 10

In general, how would you rate your physical health?
(0 = Poor, 10 = Excellent)

◯ 0 ◯ 1 ◯ 2 ◯ 3 ◯ 4 ◯ 5 ◯ 6 ◯ 7 ◯ 8 ◯ 9 ◯ 10

How would you rate your overall mental health?
(0 = Poor, 10 = Excellent )

◯ 0 ◯ 1 ◯ 2 ◯ 3 ◯ 4 ◯ 5 ◯ 6 ◯ 7 ◯ 8 ◯ 9 ◯ 10

I am content with my friendships and relationships.
(0 = Strongly Disagree, 10 = Strongly Agree )

◯ 0 ◯ 1 ◯ 2 ◯ 3 ◯ 4 ◯ 5 ◯ 6 ◯ 7 ◯ 8 ◯ 9 ◯ 10

My relationships are as satisfying as I would want them to be.
(0 = Strongly Disagree, 10 = Strongly Agree )

◯ 0 ◯ 1 ◯ 2 ◯ 3 ◯ 4 ◯ 5 ◯ 6 ◯ 7 ◯ 8 ◯ 9 ◯ 10

How often do you worry about being able to meet normal monthly living expenses?
(0 = Worry All of the Time, 10 = Do Not Ever Worry)

◯ 0 ◯ 1 ◯ 2 ◯ 3 ◯ 4 ◯ 5 ◯ 6 ◯ 7 ◯ 8 ◯ 9 ◯ 10

How often do you worry about safety, food, or housing?
(0 = Worry All of the Time, 10 = Do Not Ever Worry)

◯ 0 ◯ 1 ◯ 2 ◯ 3 ◯ 4 ◯ 5 ◯ 6 ◯ 7 ◯ 8 ◯ 9 ◯ 10

Overall Study Experience

Please rate your overall experience in the study on a scale from 1 to 5 with 1 being poor and 5 being excellent

◯ 1 ◯ 2 ◯ 3 ◯ 4 ◯ 5

On a scale of 1 to 5 (5 being very easy and 1 being very challenging), how easy was it for you to do the blood/nasal sampling and surveys with your daily routine and other responsibilities (like work or caring for others)?

○ 1 ○ 2 ○ 3 ○ 4 ○ 5

| | |
|---|---|
| Is there anything we can do to make it easier for you in the future? | ○ Yes<br>○ No |
| Please describe how we can make this process easier for future participants: | ______________________ |
| Have you participated in research studies previously? | ○ Yes<br>○ No |
| Were they remote/at-home like this study or in person studies (requiring visiting a hospital, clinic, or research site)? | ☐ Remote/at-home study<br>☐ In person study<br>(can select both if applicable) |
| Please describe the nature of the study (survey only, survey + blood/urine/other sample) | ______________________ |
| Based your experience in this study, would you participate in a similar remote or at-home study in the future? | ○ Yes<br>○ No |
| Do you think there will be any challenges for you to participate in similar studies in the future? | ○ Yes<br>○ No |
| If so, please briefly describe these challenges: | ______________________ |
| Would you participate in an in-person study requiring visiting a hospital, clinic, or research site in the future? | ○ Yes<br>○ No |
| If so, do you foresee any challenges to participating in an in-person study? | ○ Yes<br>○ No |
| If so, please briefly describe these challenges: | ______________________ |
| Receiving the Kit | |
| Did you receive your kit by the expected time? | ○ Yes<br>○ No |
| If No, Please Explain (optional). | |
| Was there any visible damage to the box or components? | ○ Yes<br>○ No |

| | |
|---|---|
| If Yes, can you describe the damage(s)? (optional) | __________________________________ |
| Did you experience any issues in receiving your kit? | ◯ Yes<br>◯ No |
| If Yes, can you describe the issue(s)? (optional) | __________________________________ |

#### Using the Kit

**How easy was it to use the homeRNA kit overall?**

| | Very easy | Somewhat easy | Neither easy nor hard | Somewhat hard | Very hard |
|---|---|---|---|---|---|
| Tasso-SST device | ○ | ○ | ○ | ○ | ○ |
| Stabilization tube | ○ | ○ | ○ | ○ | ○ |

#### How easy was it to use the Nasal swab kit?

| | Very easy | Somewhat easy | Neither easy nor hard | Somewhat hard | Very hard |
|---|---|---|---|---|---|
| Swab collection | ◯ | ◯ | ◯ | ◯ | ◯ |
| Transport tube | ◯ | ◯ | ◯ | ◯ | ◯ |

Can you please describe any part of the Instructions for use that was confusing or challenging to understand? (optional)

Returning the Kits

Were you able to use the kits and complete the collections within the designated time window?

◯ Yes
◯ No

If No, please describe what can be improved to allow you to use the kits within the designated time window.

Did you have problem(s) packaging the kit components?

◯ Yes
◯ No

If Yes, please describe the problem(s) you had.

Were the instructions for packaging and returning the kits clear?

◯ Yes
◯ No

If No, please describe what can be improved.

Did you have problem(s) filling out or accessing this survey?

◯ Yes
◯ No

If Yes, please describe the problem(s) you had.

Please use this space for any other comments on using and returning the kits.

Closing Remarks

| | |
|---|---|
| Is there anything else you would like to share with us about your experience in the study? | ◯ Yes<br>◯ No |
| Please use the space provided to share your feedback here: | ______________________ |

### Monthly Check-in

#### Sampling Survey

Please select all listed symptom(s) below that you are experiencing today.

Nose Symptoms
Runny nose ______
Congestion ______
post nasal drip ______
Sinus pain ______
Sneezing ______
Ears Symptoms
Ear pain ______
Throat Symptoms
Sore throat ______
Hoarseness ______
Chest Symptoms
Watery or itchy eyes ______
Cough ______
Phlegm or mucous production ______
Wheezing or chest tightness ______
Shortness of breath ______
Chest pain ______
Racing heart/palpitations/arrhythmia ______
Gastrointestinal Symptoms
Diarrhea ______
Nausea ______
Abdominal pain ______
Vomiting ______
Constipation ______
Reflux/GERD/heartburn ______
General Symptoms
Fatigue ______
Feeling fatigued after exercise ______
Dizziness ______
Fever ______
Chills ______
Headache ______
Aching muscles ______
Joint pain ______
Sleep Changes Symptoms
Sleep disruption/insomnia ______
Sensory Changes Symptoms
Change in or loss of smell ______
Change in or loss of taste ______
Mental Health Symptoms
Anxiety ______
Depression ______
Stress ______

Have your medications changed since you last filled out a survey?

- ☐ I started taking a new medication
- ☐ I stopped taking a medication
- ☐ My medications have not changed since I last filled out a survey

Please describe the new medication:

__________________________________

| | |
|---|---|
| Please describe which medication you stopped taking: | __________________________________________ |
| Have you received any vaccinations in the past month? | ◯ Yes<br>◯ No |
| Please select from the following list the vaccine(s) you received in the past month (select all that apply): | ☐ Flu<br>☐ COVID-19<br>☐ Other (please specify) |
| Please specify the type of vaccine: | __________________________________ |
| When did you receive this vaccine? | __________________________________ |
| When did you receive the flu vaccine? | __________________________________ |
| When did you receive the COVID-19 vaccine? | __________________________________ |
| Since our last check-in, have you tested positive for COVID-19 again? | ◯ Yes<br>◯ No |
| If you remember, when did you test positive? | __________________________________ |
| Since our last check-in, have you experienced any other (non-COVID-19) illness or health issues? | ◯ Yes<br>◯ No |
| Please specify: | __________________________________________ |

**Please respond to the following questions on a scale from 0 to 10**

Overall, how satisfied are you with life as a whole these days?
(0 = Not Satisfied at All, 10 = Completely Satisfied )

◯ 0 ◯ 1 ◯ 2 ◯ 3 ◯ 4 ◯ 5 ◯ 6 ◯ 7 ◯ 8 ◯ 9 ◯ 10

In general, how happy or unhappy do you usually feel?
(0 = Extremely Unhappy, 10 = Extremely Happy)

◯ 0 ◯ 1 ◯ 2 ◯ 3 ◯ 4 ◯ 5 ◯ 6 ◯ 7 ◯ 8 ◯ 9 ◯ 10

In general, how would you rate your physical health?
(0 = Poor, 10 = Excellent)

◯ 0 ◯ 1 ◯ 2 ◯ 3 ◯ 4 ◯ 5 ◯ 6 ◯ 7 ◯ 8 ◯ 9 ◯ 10

How would you rate your overall mental health?
(0 = Poor, 10 = Excellent )

◯ 0 ◯ 1 ◯ 2 ◯ 3 ◯ 4 ◯ 5 ◯ 6 ◯ 7 ◯ 8 ◯ 9 ◯ 10

I am content with my friendships and relationships.
(0 = Strongly Disagree, 10 = Strongly Agree )

◯ 0 ◯ 1 ◯ 2 ◯ 3 ◯ 4 ◯ 5 ◯ 6 ◯ 7 ◯ 8 ◯ 9 ◯ 10

My relationships are as satisfying as I would want them to be.
(0 = Strongly Disagree, 10 = Strongly Agree )

◯ 0 ◯ 1 ◯ 2 ◯ 3 ◯ 4 ◯ 5 ◯ 6 ◯ 7 ◯ 8 ◯ 9 ◯ 10

How often do you worry about being able to meet normal monthly living expenses?
(0 = Worry All of the Time, 10 = Do Not Ever Worry)

◯ 0 ◯ 1 ◯ 2 ◯ 3 ◯ 4 ◯ 5 ◯ 6 ◯ 7 ◯ 8 ◯ 9 ◯ 10

How often do you worry about safety, food, or housing?
(0 = Worry All of the Time, 10 = Do Not Ever Worry)

◯ 0 ◯ 1 ◯ 2 ◯ 3 ◯ 4 ◯ 5 ◯ 6 ◯ 7 ◯ 8 ◯ 9 ◯ 10

Hi [prefname],

Please read and sign this consent form.

You may open the survey in your web browser by clicking the link below:
[survey-link]

If the link above does not work, try copying the link below into your web browser:
[survey-url]

This link is unique to you and should not be forwarded to others.

Hi [prefname],

Please take this welcome and mailing address survey.

You may open the survey in your web browser by clicking the link below:
[survey-link]

If the link above does not work, try copying the link below into your web browser:
[survey-url]

This link is unique to you and should not be forwarded to others.

Thank you!

Hi [prefname],

Today will be your first sampling. You will use materials for **Timepoint 1**.

Before the blood draw, please take a look at this instructional video that guides you through the process:
https://www.youtube.com/watch?v=iV3GZ8SmmuM&embeds_euri=https%3A%2F%2Fuwbcmecovid.com%2F&feature=emb_logo

For the nasal swab sample, please follow the instructions provided in the welcome packet.

There will be **two blood draws and one nasal swab.** Most of the materials will be in the boxes labeled *Timepoint 1* but some will be in the blue boxes labled *extra supplies*.

**Please note that one sample is stabilized one is not**. This means that one blood draw will be capped immediately (the one from the white and red Tasso-SST box) and one will be attached to the clear tube with liquid inside of it (the one from the white and purple Home Blood Collection and Stabilization Kit box). **Please see the images below.**

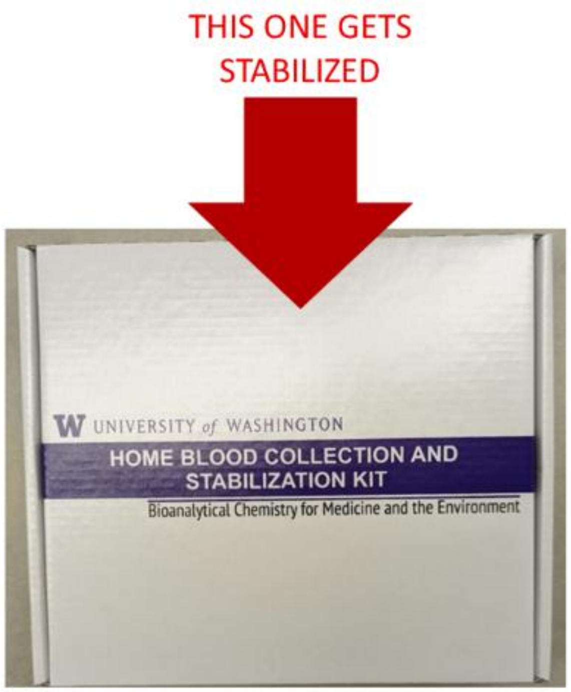


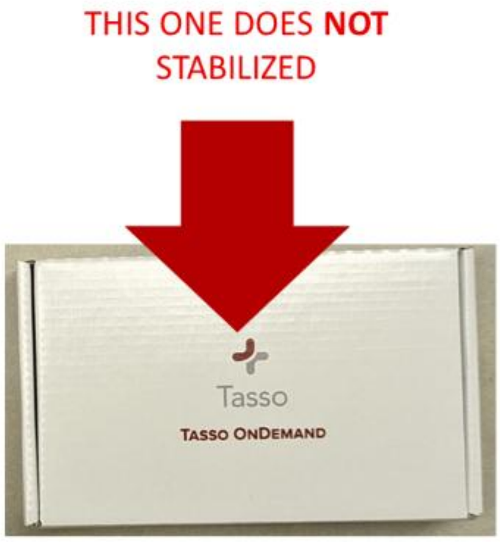

Once you are done sampling, **all samples will go into one specimen transport bag**. This bag gets sealed and placed back into the *home*RNA box (white and purple) before being put into the UPS bag and **put out for pickup the following morning (by 9:00am). If the next day is Sunday, please leave the package inside until Monday morning.**

Note: there is nothing you need to do to schedule the pickup, we automatically schedule them after you complete the surveys below.

Please complete this survey while collecting sample #1.

You may open the survey in your web browser by clicking the link below:
[survey-link]

If the link above does not work, try copying the link below into your web browser:
[survey-url]

This link is unique to you and should not be forwarded to others.

Thank you again for your participation and please let us know if you have questions or need assistance!

Sincerely,

BCME Long Covid Team

Hi [prefname],

Today you will follow the instructions for **Timepoint 2**.

There will be **one blood draw and one nasal swab.** All the materials you will need for today's sampling are in the Home Blood Collection and Stabilization Kit box (white and purple) labeled *Timepoint 2*.

**Please note that the blood sample is stabilized.** This means that it will be attached to the clear tube with the purple lid and liquid inside of it and shaken. **Please see the image below.**

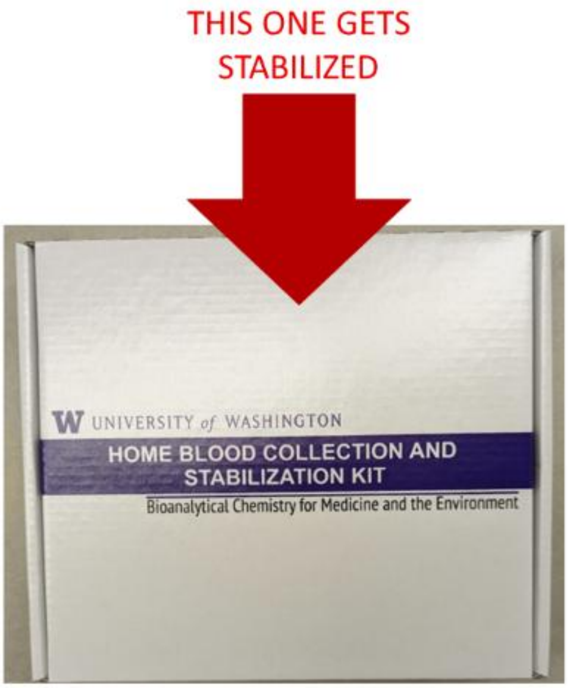


Once you are done sampling, **all samples will go into one specimen transport bag**. This bag gets sealed and placed back into the *home*RNA box (white and purple) before being put into the UPS bag and **put out for pickup the following morning (by 9:00am). If the next day is Sunday, please leave the package inside until Monday morning.**

Note: there is nothing you need to do to schedule the pickup, we automatically schedule them after you complete the surveys below.

Please complete this survey while collecting sample #2.

You may open the survey in your web browser by clicking the link below:
[survey-link]

If the link above does not work, try copying the link below into your web browser:
[survey-url]

This link is unique to you and should not be forwarded to others.

Sincerely,

BCME Long Covid Team

Hi [prefname],

Today will be your first extended sampling. You will follow the instructions for **Timepoint 6**.

There will be **one blood draw and one nasal swab.** All the materials you will need for today's sampling are in the Home Blood Collection and Stabilization Kit box (white and purple) labeled *Timepoint 2*.

**Please note that the blood sample is stabilized.** This means that it will be attached to the clear tube with the purple lid and liquid inside of it and shaken. **Please see the image below.**

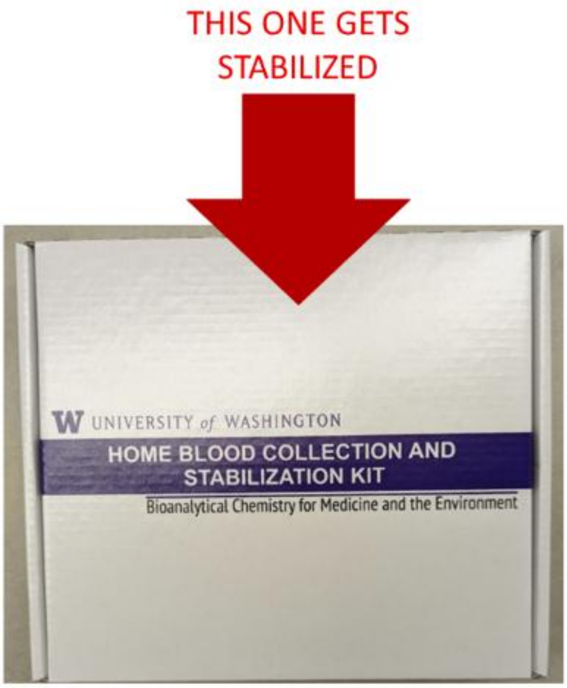


Once you are done sampling, **all samples will go into one specimen transport bag**. This bag gets sealed and placed back into the *home*RNA box (white and purple) before being put into the UPS bag and **put out for pickup the following morning (by 9:00am). If the next day is Sunday, please leave the package inside until Monday morning.**

Note: there is nothing you need to do to schedule the pickup, we automatically schedule them after you complete the surveys below.

Please complete this survey while collecting sample #6.

You may open the survey in your web browser by clicking the link below:
[survey-link]

If the link above does not work, try copying the link below into your web browser:
[survey-url]

This link is unique to you and should not be forwarded to others.

Sincerely,

BCME Long Covid Team

Hi [prefname],

It has been 4 months since you submitted your first survey and sample.

We are asking you to complete 3 additional follow-up surveys. We are asking you to complete the first one now, which is 4 months after your initial participation. The next one will be one more from now (month 5) and the last one will be two months from now (month 6). This information will be combined with your samples and survey data previously collected. If you have any questions about this survey, please contact the study team at 206-295-9238. You will be paid a $20 gift card for completing each additional survey.

You may open the survey in your web browser by clicking the link below:
[survey-link]

If the link above does not work, try copying the link below into your web browser:
[survey-url]

This link is unique to you and should not be forwarded to others.

Thank you!

### SWAB INSTRUCTIONS

*Self-swab nasal speciman collection*

#### KIT CONTENTS

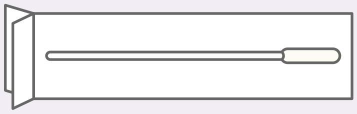

**1 x STERILE SWAB**

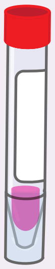

**1 x COLLECTION TUBE**

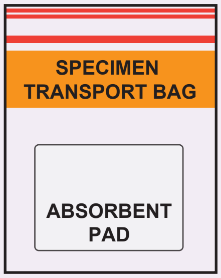


**1 x SPECIMEN BAG**

#### GETTING READY

1. Blow your nose. Discard the tissue.
   (Blowing your nose makes it easier to swab the inside of your nose)
2. Wash your hands with soap and water for 20 seconds - OR - use a hand sanitizer that contains at least 60% alcohol.
3. Identify a clean, dry surface to perform the nasal swab collection.
4. Open the kit and place all the supplies on the clean, dry surface.

### SWAB INSTRUCTIONS

*Self-swab nasal speciman collection*

#### COLLECTING NASAL SPECIMEN

**1**

##### LOOSEN CAP ON COLLECTION TUBE

Slightly loosen the cap and safely place the tube in a location where it won't spill.
(You will be putting the swab into this tube when finished)

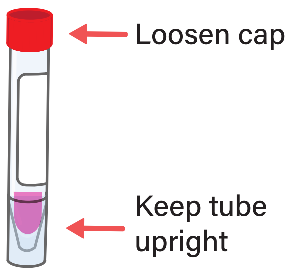


**2**

##### OPEN NASAL SWAB

Open the swab and keep it within easy reach.
(Be careful to touch ONLY the handle and NOT the tip)

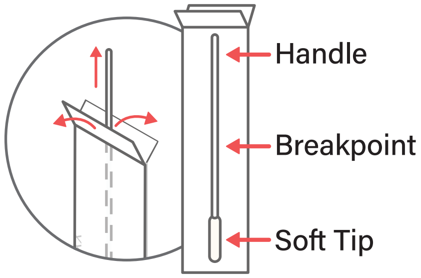


**3**

##### SWAB NOSE (BOTH NOSTRILS)

Gently insert the entire soft tip of the swab into one nostril until you feel a bit of resistance. Slowly rotate the swab in a circular motion against the inside of your nostril at least 4 times for about 15 seconds.

Be sure to collect any nasal drainage that may be present on the swab.

Next, gently insert the same swab in the other nostril and repeat the same 15-second procedure.

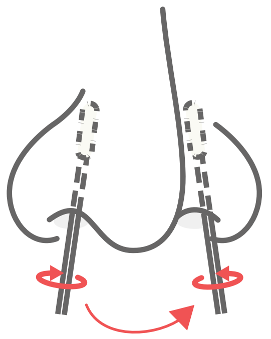

**4**

##### PLACE SWAB INTO COLLECTION TUBE

Place swab tip first into the provided tube.

Break off the swab handle at the swab breakpoint by bending back and forth.

Screw the cap back on tightly.

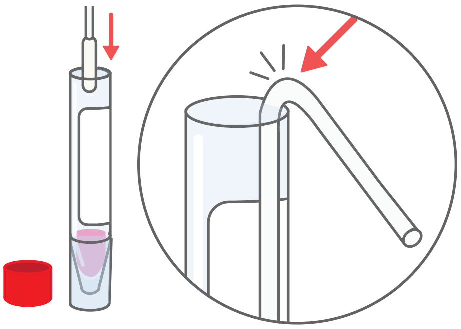

**5**

##### PLACE TUBE INTO SPECIMEN TRANSPORT BAG

Using a tissue, pick up the tube and place into the provided specimen transport bag. Discard the tissue. Seal the bag.

Refer to "Shipping Intructions" on mailing the sample back to the lab.

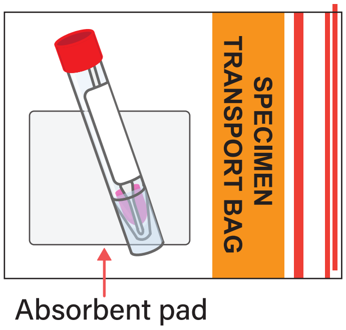

Dear **Participant**,

Thank you once again for participating in the extended sampling study!

Like in the original portion of the study, the study kit you just received contains all the necessary materials for collecting the blood samples and nasal swabs. **The boxes are labeled with the timepoints that correspond with the schedule included in the welcome packet**. Please refer to the *Sampling Schedule* for information about the when you will collect samples and what kinds of samples you will collect at each timepoint. Any additional materials you may need are included in the blue boxe labeled *Extra Supplies* along with backup materials should you encounter any issues.

In this box, you will also find pre-labeled UPS LabPaks which you will use to send the samples back to our lab. After each timepoint (once you've completed both the blood draw(s), nasal swab, and the surveys), you will place the materials into the LabPak according to the enclosed instructions and leave it at the designated location for pickup.

Your first sampling will be on **Date**. Once you are done sampling, **please leave the package outside of your door the following morning before 9:00am. If the following day is a Sunday, please wait to put your package out until Monday morning**. If the package does not get picked up or if there are issues, please reach out and let us know so that we can schedule a new pickup for the samples.

You will also receive email notifications for each sampling and survey timepoint with additional instructions. If at any point you have any questions or concerns, please don't hesitate to contact us at (XXX) XXX-XXXX or at.

Sincerely,

Filip Stefanovic

PhD Student, BCME Lab

University of Washington

#### PREPARE DEVICES AND APPLICATION SITE

FIRST TIME USERS

Point your phone's camera here to watch an instructional video:

SCAN ME

or go to https://uwbcme-covid.com/study-resources/

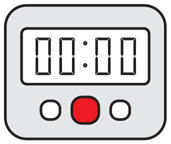


1. Wash hands and get a timer.

You will use a timer in Steps 2 and 10.

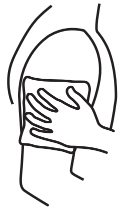

2. Apply hot pack to upper arm for 2 minutes.

Warming helps your blood flow better.

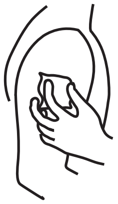

3. Clean arm with alcohol wipe.

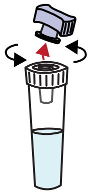

4. Open the stabilizer tube by twisting off the purple cap.

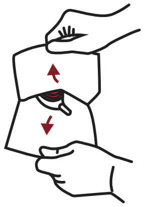

5. Open Tasso pouch by pulling apart white and clear layers.

Discard cap in pouch.

#### COLLECT BLOOD USING TASSO-SST

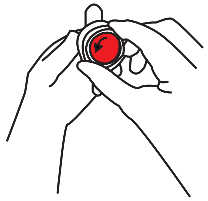

6. Remove clear plastic cover over the red button.

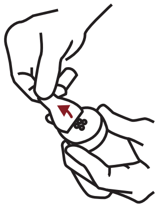

7. Peel paper tab behind the red button.

Keep the tube pointing down.

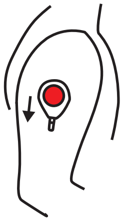

8. Stick device to shoulder.

Do not remove once it is on.

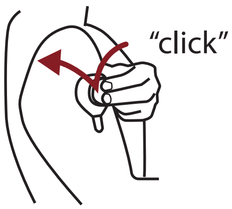


9. Press button quickly and firmly until it can't go any farther. Wait 2 seconds then let go.

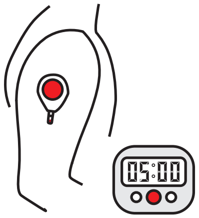


10. Start a 5 minute timer.

Keep arm at your side. You wont see blood right away. It can take up to a minute for blood to flow.

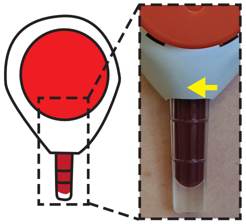

11. After 5 minutes or when the tube fills, whichever comes first, peel off the device.

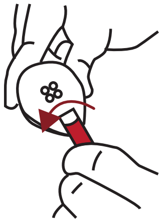

12. Remove tube by firmly twisting a quarter turn and pulling down.

This may take a bit of finger strength.

#### MIX AND PACKAGE

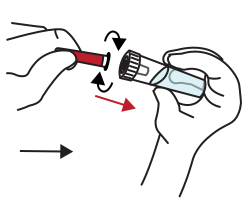

13. Bring together the blood tube and stabilizer tube and screw these together tightly.

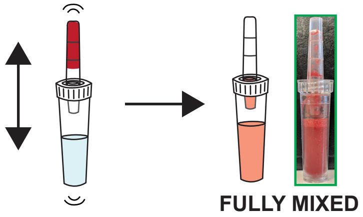


14. With the stabilizer tube on the bottom, shake hard up and down to mix. Stop when mixed.

Some fluid may remain in blood tube. When mixed, the color is the same. See insert for details.

DO NOT DISCONNECT TUBES.

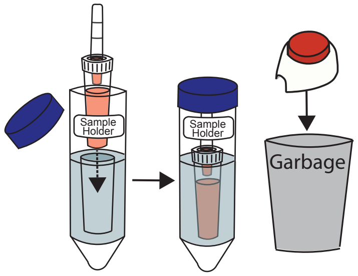


15. Place sample in sample holder. Throw away used Tasso device.

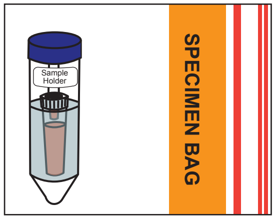


16. Place blood sample in the specimen bag.

Leave the absorbent pad in the bag.

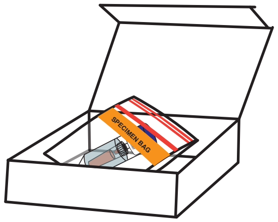

17. Place specimen bag in box and fill out collection date and time on the box.

Refer to the shipping instructions for how to mail this kit back to the lab.

### INSTRUCTIONS FOR USE

#### Home Blood Collection Kit

**Thank you for participating in our study!**

#### KIT CONTENTS

Make sure your kit contains all components listed below

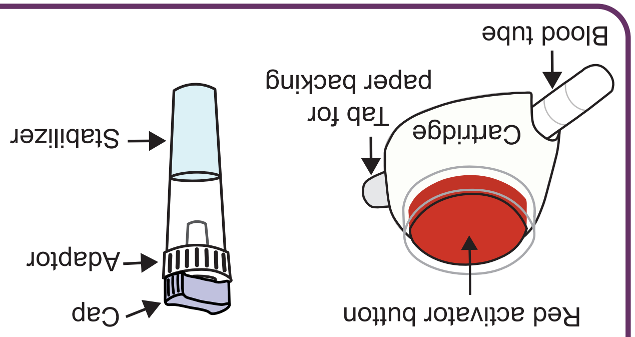


**1) Tasso-SST device**
**2) Stabilizer tube**
**3) Alcohol wipes**
**4) Hot pack**
**5) Bandage**
**6) Specimen bag**
**7) Sample Holder**
**8) Mail return bag**

#### INTENDED USE

TASSO-SST is a single use blood collection device that is intended for the self collection of capillary blood from the upper arm of adults (18 years or older). The stabilizer tube contains liquid that is intended for stabilizing the collected blood. The home blood sampling kit is for academic research use.

#### STORAGE

Store at 15 - 30ºC (60 - 80ºF) in a dry place.

#### WARNINGS

- The TASSO-SST is a sterile device. Do not open until use.
- The TASSO-SST device contains sharps. Handle with care.
- For external use only.
- Keep out of reach of children.
- Use while seated as fainting may occur during blood sampling procedure.
- Do not activate the red button until device is firmly on skin.
- Wipe application site with alcohol wipe to reduce infection risk.
- Always use a new unopened pouch of Tasso-SST. Do not re-use.

#### ONLINE SURVEY

Please check your e-mail or text message for a link to fill out the daily use online survey.

### Sampling Schedule

| | | |
|---|---|---|
| Day 1 - 4 | **Enrollment**<br>informed consent, phone call, survey, kit delivery | Date |
| Day 5 | **Timepoint 1**<br>two blood draws, nasal swab, survey | X/X/XXXX |
| Day 10 | **Timepoint 2**<br>one blood draw, nasal swab, survey | X/X/XXXX |
| Day 15 | **Timepoint 3**<br>one blood draw, nasal swab, survey | X/X/XXXX |
| Day 20 | **Timepoint 4**<br>one blood draw, nasal swab, survey | X/X/XXXX |
| Day 25 | **Timepoint 5**<br>two blood draws, nasal swab, two surveys | X/X/XXXX |

### SHIPPING INSTRUCTIONS

#### SAMPLE PACKAGING INSTRUCTIONS TIMEPOINTS 1, 5

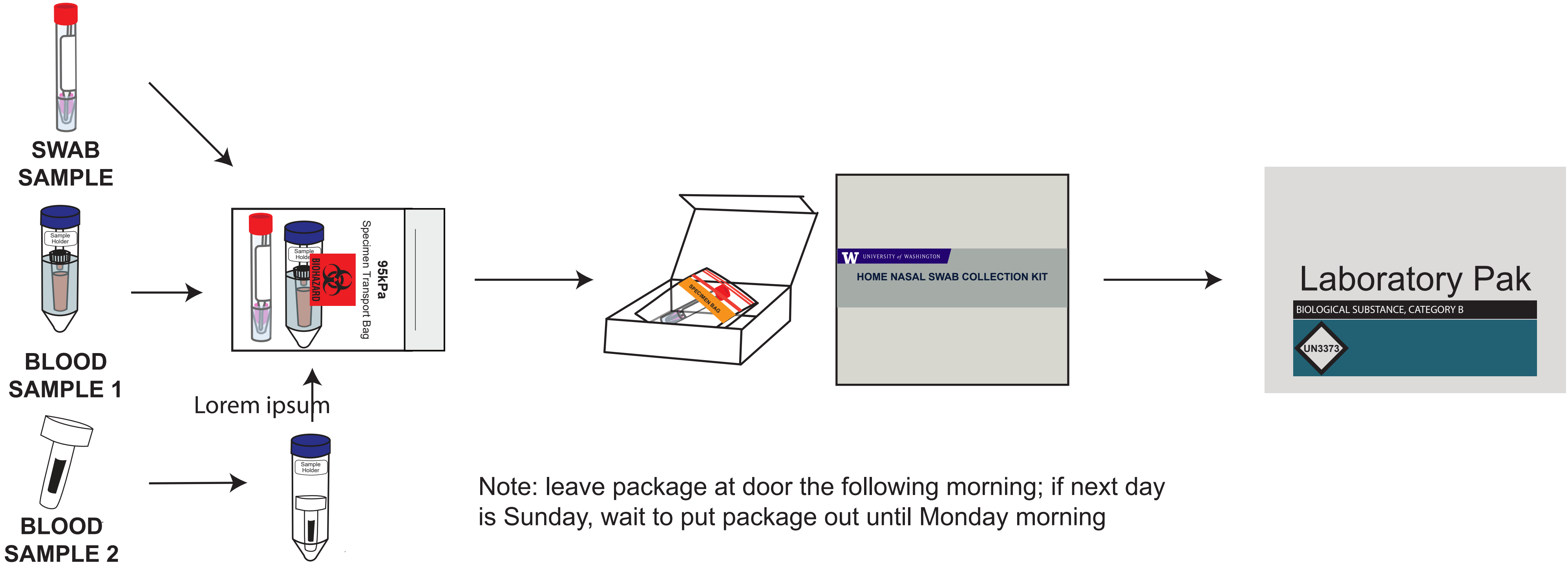


Note: leave package at door the following morning; if next day is Sunday, wait to put package out until Monday morning

#### SAMPLE PACKAGING INSTRUCTIONS TIMEPOINTS 2, 3, 4

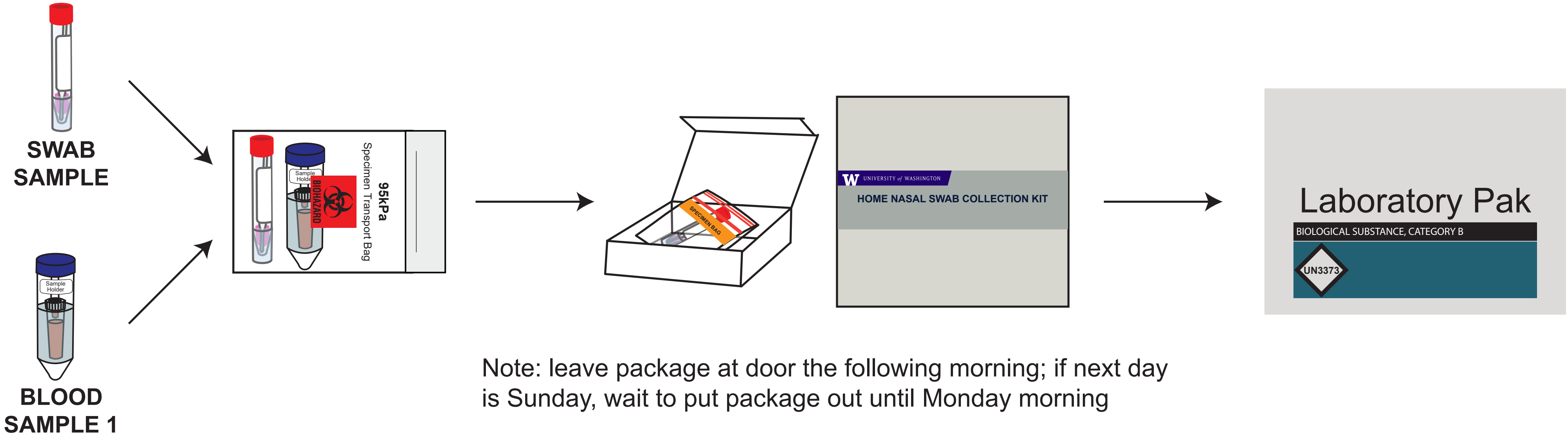


Note: leave package at door the following morning; if next day is Sunday, wait to put package out until Monday morning

### This timepoint requires **2 blood samples.**

One blood sample is stabilized with the ***home*RNA kit** labeled Timepoint 1 (1st timepoint) or Timepoint 5 (last timepoint) and is **stabilized with the stabilizer tube.**

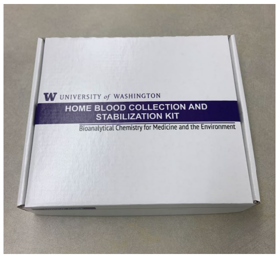


*home*RNA kit

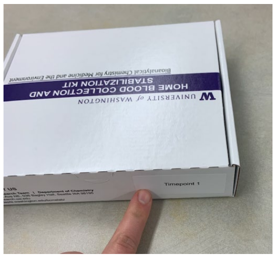

Timepoint label

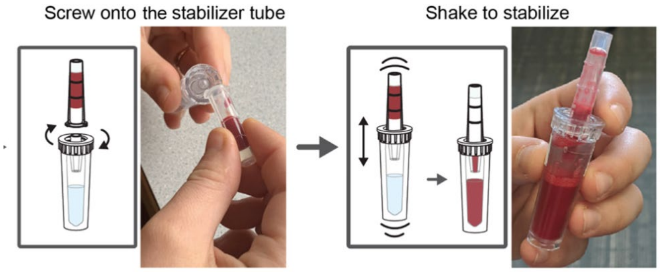


Stabilized blood sample with *home*RNA

The 2nd blood sample uses the **Tasso OnDemand box** labeled Timepoint 1 or Timepoint 5 and is **capped.**

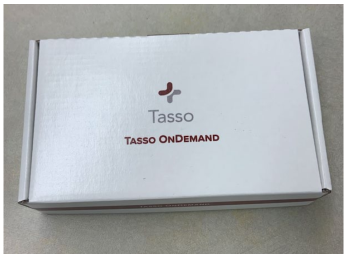


Tasso OnDemand box

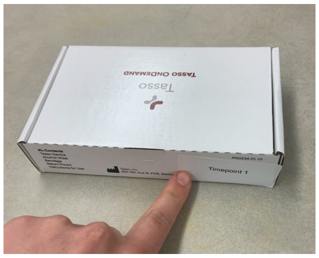

Timepoint label

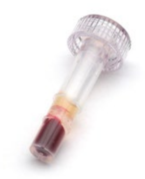

Capped blood sample

Dear Participant,

Thank you once again for contributing to our study!

The study kit you just received contains all the necessary materials for collecting the blood samples and nasal swabs. **The boxes are labeled with the timepoints that correspond with the schedule included in the welcome packet**. Please refer to the *Sampling Schedule* for information about the when you will collect samples and what kinds of samples you will collect at each timepoint.

Any additional materials you may need are included in the blue boxes labeled *Extra Supplies* along with backup materials should you encounter any issues. There is also a small white box in the corner with the temperature and humidity monitor which you will use when completing the surveys and are welcome to keep after you complete the study.

In this box, you will also find pre-labeled UPS LabPaks which you will use to send the samples back to our lab. After each timepoint (once you've completed both the blood draw(s), nasal swab, and the surveys), you will place the materials into the LabPak according to the enclosed instructions and leave it at the designated location for pickup.

If possible, **please complete the sampling for the first timepoint on the day you receive this package**. Once you are done sampling, **please leave the package outside of your door the following morning before 9:00am. If the following day is a Sunday, please wait to put your package out until Monday morning**. If the package does not get picked up or if there are issues, please reach out and let us know so that we can schedule a new pickup for the samples.

Additionally, we've included a University of Washington mug as a small token of our appreciation for participating in this study.

If at any point you have any questions or concerns, please don't hesitate to contact us at (XXX) XXX-XXXX or at.

Sincerely,

Filip Stefanovic
PhD Student, BCME Lab
University of Washington
